# Effects of a multimedia campaign on HIV self-testing and PrEP outcomes among young people in South Africa: A mixed-methods impact evaluation of ‘MTV Shuga Down South’

**DOI:** 10.1101/2021.10.16.21264987

**Authors:** Isolde Birdthistle, Sarah Mulwa, Sophie Sarrassat, Venetia Baker, David Khanyile, Dominique O’Donnell, Cherie Cawood, Simon Cousens

**Affiliations:** Faculty of Epidemiology & Population Health, London School of Hygiene & Tropical Medicine, Keppel Street, London WC1E 7HT, United Kingdom; Epicentre Health Research, Hillcrest, Durban, KwaZulu-Natal 3610, South Africa

**Keywords:** HIV prevention, adolescents, young people, media, evaluation, South Africa

## Abstract

**Introduction:** Innovative HIV technologies can help to reduce HIV incidence, yet uptake of such tools is relatively low among young people. To create awareness and demand among adolescents and young adults, a new campaign of the pan-African MTV Shuga series (“Down South 2”; DS2), featured storylines and messages about HIV self-testing (HIVST) and pre-exposure prophylaxis (PrEP) through television, radio and accompanying multimedia activities in 2019-2020.

**Methods:** We conducted a mixed-methods evaluation of the new MTV Shuga series among 15-24 year-olds in Eastern Cape, South Africa, in 2020. A web-based survey, promoted via social media platforms of schools, universities, and communities, assessed exposure to MTV Shuga and knowledge of HIV status; secondary outcomes included awareness and uptake of HIVST and PrEP. An embedded qualitative evaluation explored mechanisms of MTV Shuga’s impact through in-depth individual and group interviews.

**Results:** Among 3,431 online survey participants, 43% had engaged with MTV Shuga and 24% with DS2 specifically. Knowledge of HIV status was higher among those exposed to DS2 (58%) versus those who were not (35%; adjusted OR=2.06 [95% CI:1.64-2.58]). Exposure was also associated with increased awareness of HIVST (60% vs 28%; aOR=1.99[1.61-2.47]) and use of HIVST (29% vs 10%; aOR=2.49[1.95-3.19]). One-third of respondents were aware of PrEP, with higher proportions among those exposed versus non-exposed to DS2 (52% vs 27%; aOR=1.90[1.53-2.35]). Qualitative insights identified mechanisms by which DS2 increased awareness, confidence and motivation to use HIVST and PrEP, but had less influence on service access.

**Conclusions:** We found evidence consistent with a positive causal impact of the MTV Shuga DS2 campaign on HIV prevention outcomes among young people in a high-prevalence setting. As diverse testing and PrEP technologies become accessible, an immersive edutainment campaign can help to expand HIV prevention choices and close age and gender gaps in HIV testing and prevention goals.

## Introduction

Biomedical innovations in HIV prevention are expanding the options and tools to avoid HIV acquisition and accelerate declines in HIV incidence. By diversifying HIV testing choices, HIV self-test (HIVST) kits have enabled more people to know their HIV status and access pathways into treatment and prevention.[1] Confirmation of positive status can link those living with HIV/AIDS to life-saving treatment, viral suppression and improved health, while reducing onward transmission.[2] Knowledge of negative HIV status can link individuals to prevention services, including highly efficacious pre-exposure prophylaxis (PrEP), to remain uninfected.

New tools like HIVST and PrEP can be particularly valuable for young people, an important demographic group for HIV epidemic control and the least likely to know their HIV status.[2] As yet, no country in sub-Saharan Africa has achieved the first step of the UNAIDS “90:90:90” targets for the HIV treatment cascade (90% knowledge of status) among 15-24 year-olds, despite significant gains in diagnostic coverage overall.[3] This represents a missed opportunity for early diagnosis to avoid illness and transmission, and for more young people to avail themselves of high-impact prevention options like PrEP.[4, 5]

When offered HIV self-testing in the context of research studies, young people have welcomed the opportunity to test in private, without the judgment, stigma or discrimination they often face in facility-based services.[6-8] In real-world conditions, relatively few young people are aware of or utilising HIVST.[9] Similarly, while research has shown PrEP to be an acceptable HIV prevention method for young men and women, awareness and uptake of PrEP via national programmes is lower than expected, and annual growth has slowed over time since 2016.[10-13] Identifying ways to expand the reach and engagement of young people in HIV prevention, particularly testing services and PrEP, has become a priority for HIV epidemic control.[1, 3]

MTV Shuga is a multimedia campaign based around a popular dramatic series that promotes HIV prevention and positive sexual health messaging with entertaining storylines and characters. Since the first series based in Nairobi in 2009, MTV Shuga has been broadcast on 179 terrestrial channels reaching an estimated 719 million households. All episodes are available rights-free on internet platforms and MTV Staying Alive Foundation estimates 42 million people have been reached through social media.[14] In 2019-2020, a new series entitled ‘Down South 2’ was produced in South Africa and incorporated storylines designed to increase awareness and demand for HIV testing, including self-testing, and PrEP in ways that resonate with young people. The show was complemented by wrap-around, “360-media” activities including a radio series, documentary films, and online resources.[14] (Figure S1) In the town of Mthatha in Eastern Cape, South Africa, peer-education and community events and a graphic novel distributed through schools enabled opportunities for offline engagement among those underserved by television and internet.

To date, the efficacy of MTV Shuga has been demonstrated in a cluster-randomised trial of community viewings in Nigeria, which showed a positive impact on HIV and STI testing.[15] However, evidence of effectiveness in non-trial conditions is limited.[16, 17] We sought to evaluate the impact of MTV Shuga Down South on HIV prevention outcomes, including awareness and uptake of HIVST and PrEP, among young people in South Africa.

## Methods

A mixed-methods evaluation was conducted in 2020 to determine whether and how MTV Shuga Down South 2 (DS2) impacted HIV prevention outcomes among young people. All research activities were conducted remotely to avoid risk of SARS-CoV-2 transmission.

First, an online survey was hosted on a website, free of charge for users through reverse-charging arrangements with the service provider. Those who completed the survey received mobile data credit of SAR50 transferred to a phone number provided for this purpose only. The survey was open to 15-24 year-old males and females and promoted through virtual marketing on Facebook, Instagram, and social media platforms of schools, universities, community groups and clinics in Mthatha, Eastern Cape. This setting was targeted due to: recent distribution of HIV self-testing kits and availability of oral PrEP (ensuring a supply to meet any demand generated by the DS2 campaign);[18] availability of the offline components of the DS2 campaign, described above; less HIV prevention research and lower HIV testing levels in Eastern Cape relative to other provinces; and the recommendation of the South African Department of Health.

The self-administered questionnaire (File S1) was anonymised with no name or other personal identifying information requested and questions designed to measure exposure to the MTV Shuga DS2 campaign (primary exposure) and any other MTV Shuga campaign (secondary exposure). The primary outcome was knowledge of HIV status, i.e., the proportion who tested for HIV in the past year and received the result, or ever tested HIV positive. Secondary outcomes included awareness, willingness to use, and uptake of HIVST and PrEP.

We constructed a directed acyclic graph (DAG) to represent the hypothesised causal relationship between intervention exposure, the study outcomes, and other socio-demographic characteristics.[19] (Figure S2) The DAG was interrogated to identify the minimal set of constructs needed to control for confounding. We calculated the proportion of respondents who knew their HIV status, and each secondary outcome, by exposure to DS2, and estimated associations between the intervention and outcomes using multivariable logistic regression to adjust for confounding variables informed by the DAG. Interaction terms were included where there was evidence of effect modification by age or gender. Each logistic regression model was restricted to individuals with non-missing responses for each outcome of interest.

A sample of participants who reportedly engaged with DS2 (watched or listened to at least one episode on TV, internet or radio) and opted into further research were invited to participate in an embedded qualitative evaluation to explore mechanisms of MTV Shuga’s impact. Qualitative research activities included in-depth interviews (18 female, 13 male) and six group discussions, held in age- and gender-specific groups of 4-6 participants. Topic guides (Files S2, S3) were based on the ‘COM-B’ behavioural wheel model to explore MTV Shuga DS2’s influence on participants’ capability, opportunity, and motivation to adopt attitudes and behaviours supportive of HIVST and PrEP.[20] Video clips with DS2 scenes about HIVST and PrEP were shown to generate discussion. Trained researchers who were bilingual (in either isiXhosa or Zulu and English), and aged under 30 years, facilitated the individual and group interviews via phone, Zoom or WhatsApp. Participants received data transfers in advance, to facilitate participation, and SAR100 airtime credit upon completion. Researchers transcribed all interviews into English and transcripts were analysed using a hybrid (deductive and inductive) thematic coding process.[21] Deductive codes were generated around the three conditions of the COM-B behavioural framework. A stage of open coding was conducted to allow other mechanisms of influence to emerge inductively.

### Research Ethics Statement

Ethics approvals were received by the University of KwaZulu-Natal, London School of Hygiene & Tropical Medicine, and the World Health Organisation. Participants provided online consent and parents or guardians provided consent for participants under 18 years.

### Patient and Public Involvement

Members of the public are engaged in the dissemination and discussion of results, via social media platforms and a public webinar on MTV Shuga evidence.

## Results

### Online Survey

#### Characteristics of the survey sample

The web-based survey was available online from September to December 2020, during which 4,145 records were created. After removing records without full consent (n=407) or gender (n=144), and likely duplicates (n=163), 3,431 (83%) records were taken forward for analysis.(Table 1) Of those, respondents were predominantly female (59%), aged 20-24 years (69% versus 31% aged 15-19), and spoke IsiXhosa at home (80%). Most respondents (83%) were enrolled in education, including 34% in university, 28% in technical/vocational college, and 21% in primary or secondary school, while 3.1% were employed and 10.4% unemployed. The majority resided in urban settings (85%), primarily in Mthatha town (72%) or elsewhere in Eastern Cape province (11%), while 16% lived in other provinces of South Africa (6%, 4% and 3% in Western Cape, Gauteng, and KwaZulu-Natal, respectively). About 40% of participants reported experiencing food insecurity (going to bed hungry in the past month) either sometimes, often or always.

**Table 1.** Socio-demographic characteristics of online survey participants, overall and by age group (N=3,431)

|  | Age group |  |  |  |  |  |
| --- | --- | --- | --- | --- | --- | --- |
|  | 15-19 years<br>(N=1079) |  | 20-24 years<br>(N=2352) |  | Total<br>(N=3431) |  |
|  | n | % | n | % | n | % |
| ***Socio-demographic characteristics*** |  |  |  |  |  |  |
| **Gender** |  |  |  |  |  |  |
| Male | 372 | 34.5 | 945 | 40.2 | 1,317 | 38.4 |
| Female | 656 | 60.8 | 1,364 | 58.0 | 2,020 | 58.9 |
| Other | 51 | 4.7 | 43 | 1.8 | 94 | 2.7 |
| **Current schooling/employment status** |  |  |  |  |  |  |
| In school (primary/secondary) | 597 | 55.3 | 129 | 5.5 | 726 | 21.2 |
| TVET<sup>†</sup> | 144 | 13.3 | 823 | 35.0 | 967 | 28.2 |
| University | 228 | 21.1 | 936 | 39.8 | 1,164 | 33.9 |
| Employed | 12 | 1.1 | 94 | 4.0 | 106 | 3.1 |
| Unemployed | 49 | 4.5 | 308 | 13.1 | 357 | 10.4 |
| Unknown | 49 | 4.5 | 62 | 2.6 | 111 | 3.2 |
| **Language spoken at home** |  |  |  |  |  |  |
| English | 99 | 9.2 | 160 | 6.8 | 259 | 7.5 |
| IsiXhosa | 798 | 74.0 | 1,947 | 82.8 | 2,745 | 80.0 |
| Zulu | 100 | 9.3 | 124 | 5.3 | 224 | 6.5 |
| Other | 72 | 6.7 | 119 | 5.1 | 191 | 5.6 |
| Unknown | 10 | 0.9 | 2 | 0.1 | 12 | 0.3 |
| **Province** |  |  |  |  |  |  |
| Eastern Cape (EC) - Mthatha | 716 | 66.4 | 1,746 | 74.2 | 2,462 | 71.8 |
| Eastern Cape - O.R. Tambo | 24 | 2.2 | 46 | 2.0 | 70 | 2.0 |
| Eastern Cape - other EC | 102 | 9.5 | 192 | 8.2 | 294 | 8.6 |
| Western Cape | 57 | 5.3 | 157 | 6.7 | 214 | 6.2 |
| Kwa-Zulu Natal | 65 | 6.0 | 38 | 1.6 | 103 | 3.0 |
| Gauteng | 56 | 5.2 | 80 | 3.4 | 136 | 4.0 |
| Other provinces | 36 | 3.3 | 47 | 2.0 | 83 | 2.4 |
| Unknown | 23 | 2.1 | 46 | 2.0 | 69 | 2.0 |
| **Urban/Rural residence<sup>‡</sup>** |  |  |  |  |  |  |
| Urban setting | 891 | 82.6 | 2,032 | 86.4 | 2,923 | 85.2 |
| Rural setting | 95 | 8.8 | 192 | 8.2 | 287 | 8.4 |
| Unknown | 93 | 8.6 | 128 | 5.4 | 221 | 6.4 |
| **Food insecurity** |  |  |  |  |  |  |
| Never/rarely | 610 | 56.5 | 1,268 | 53.9 | 1,878 | 54.7 |
| Sometimes | 319 | 29.6 | 820 | 34.9 | 1,139 | 33.2 |
| Often/always | 72 | 6.7 | 146 | 6.2 | 218 | 6.4 |
| Unknown | 78 | 7.2 | 118 | 5.0 | 196 | 5.7 |
| **Called a helpline or searched for information on HIV on the internet<sup>§</sup>** |  |  |  |  |  |  |
| No | 485 | 44.9 | 1,182 | 50.3 | 1,667 | 48.6 |
| Yes | 422 | 39.1 | 930 | 39.5 | 1,352 | 39.4 |
| Unknown | 172 | 15.9 | 240 | 10.2 | 412 | 12.0 |

<sup>†</sup> TVET: Technical Vocational Educational and Training colleges
<sup>‡</sup> coding Mthatha to urban setting; <sup>§</sup> this measure includes the following sites/helplines: Bwise, Loveline, Childline, but excludes MTV Shuga website

Most participants lived in a household with a television (83%), internet (75%), radio (72%), and a computer or other digital device (57%); household ownership of all these was higher among older (20-24 year-olds) than younger respondents (15-19 year-olds).(Table 2) Most respondents owned their own smartphone (85% for both age groups), while about half owned their own computer and fewer (35%) owned a radio. Digital media engagement was high with 86% using the internet and social media platforms at least once a week, most commonly Facebook (72%), YouTube (44%), and Instagram (37%). Fewer than 1% reported never using the internet or social media. Most watched TV (74%) and listened to the radio (62%) at least weekly. Use of all media types was higher among older than younger respondents, although data were more often missing among the younger group.

**Table 2.** Media ownership and sexual experiences reported by online survey participants, overall and by age group and gender (N=3,431)

|  | 15-19 years |  |  |  |  |  |  |  | 20-24 years |  |  |  |  |  |  |  |
| --- | --- | --- | --- | --- | --- | --- | --- | --- | --- | --- | --- | --- | --- | --- | --- | --- |
|  | Male<br>(N=372) |  | Female<br>(N=656) |  | Other<br>(N=51) |  | Total<br>(N=1,079) |  | Male<br>(N=945) |  | Female<br>(N=1,364) |  | Other<br>(N=43) |  | Total<br>(N=2,352) |  |
|  | n | % | n | % | n | % | n | % | n | % | n | % | n | % | n | % |
| **Media assets** |  |  |  |  |  |  |  |  |  |  |  |  |  |  |  |  |
| **Household ownership** |  |  |  |  |  |  |  |  |  |  |  |  |  |  |  |  |
| Radio | 240 | 64.5 | 463 | 70.6 | 17 | 33.3 | 720 | 66.7 | 727 | 76.9 | 1,002 | 73.5 | 15 | 34.9 | 1,744 | 74.1 |
| Television (TV) | 291 | 78.2 | 533 | 81.3 | 19 | 37.3 | 843 | 78.1 | 811 | 85.8 | 1,160 | 85 | 18 | 41.9 | 1,989 | 84.6 |
| Computer / device | 200 | 53.8 | 334 | 50.9 | 11 | 21.6 | 545 | 50.5 | 589 | 62.3 | 801 | 58.7 | 14 | 32.6 | 1,404 | 59.7 |
| Internet | 270 | 72.6 | 464 | 70.7 | 18 | 35.3 | 752 | 69.7 | 752 | 79.6 | 1,038 | 76.1 | 20 | 46.5 | 1,810 | 77.0 |
| TV subscription | 184 | 49.5 | 352 | 53.7 | 10 | 19.6 | 546 | 50.6 | 428 | 45.3 | 657 | 48.2 | 9 | 20.9 | 1,094 | 46.5 |
| Unknown | 33 | 8.9 | 51 | 7.8 | 1 | 2.0 | 85 | 7.9 | 47 | 5.0 | 84 | 6.2 | 2 | 4.7 | 133 | 5.7 |
| Household media assets index |  |  |  |  |  |  |  |  |  |  |  |  |  |  |  |  |
| Low | 125 | 33.6 | 245 | 37.3 | 40 | 78.4 | 410 | 38 | 294 | 31.1 | 453 | 33.2 | 26 | 60.5 | 773 | 32.9 |
| Medium | 111 | 29.8 | 168 | 25.6 | 2 | 3.9 | 281 | 26 | 330 | 34.9 | 433 | 31.7 | 8 | 18.6 | 771 | 32.8 |
| High | 103 | 27.7 | 192 | 29.3 | 8 | 15.7 | 303 | 28.1 | 274 | 29.0 | 394 | 28.9 | 7 | 16.3 | 675 | 28.7 |
| Unknown | 33 | 8.9 | 51 | 7.8 | 1 | 2.0 | 85 | 7.9 | 47 | 5.0 | 84 | 6.2 | 2 | 4.7 | 133 | 5.7 |
| **Personal ownership** |  |  |  |  |  |  |  |  |  |  |  |  |  |  |  |  |
| Radio | 133 | 35.8 | 215 | 32.8 | 3 | 5.9 | 351 | 32.5 | 373 | 39.5 | 455 | 33.4 | 10 | 23.3 | 838 | 35.6 |
| Smartphone | 322 | 86.6 | 573 | 87.3 | 18 | 35.3 | 913 | 84.6 | 813 | 86.0 | 1,179 | 86.4 | 17 | 39.5 | 2,009 | 85.4 |
| Computer / device | 166 | 44.6 | 265 | 40.4 | 6 | 11.8 | 437 | 40.5 | 540 | 57.1 | 755 | 55.4 | 12 | 27.9 | 1,307 | 55.6 |
| Unknown | 32 | 8.6 | 51 | 7.8 | 1 | 2.0 | 84 | 7.8 | 899 | 95.1 | 1,279 | 93.8 | 41 | 95.3 | 2,219 | 94.3 |
| Individual media assets index |  |  |  |  |  |  |  |  |  |  |  |  |  |  |  |  |
| Low | 133 | 35.8 | 249 | 38 | 44 | 86.3 | 426 | 39.5 | 291 | 30.8 | 453 | 33.2 | 27 | 62.8 | 771 | 32.8 |
| Medium | 125 | 33.6 | 246 | 37.5 | 5 | 9.8 | 376 | 34.8 | 342 | 36.2 | 500 | 36.7 | 6 | 14 | 848 | 36.1 |
| High | 82 | 22 | 110 | 16.8 | 1 | 2.0 | 193 | 17.9 | 266 | 28.1 | 326 | 23.9 | 8 | 18.6 | 600 | 25.5 |
| Unknown | 32 | 8.6 | 51 | 7.8 | 1 | 2.0 | 84 | 7.8 | 46 | 4.9 | 85 | 6.2 | 2 | 4.7 | 133 | 5.7 |
| How often do you watch TV |  |  |  |  |  |  |  |  |  |  |  |  |  |  |  |  |
| Never | 8 | 2.2 | 17 | 2.6 | 3 | 5.9 | 28 | 2.6 | 21 | 2.2 | 20 | 1.5 | 3 | 7 | 44 | 1.9 |
| Less than weekly | 46 | 12.4 | 105 | 16 | 6 | 11.8 | 157 | 14.6 | 125 | 13.2 | 169 | 12.4 | 4 | 9.3 | 298 | 12.7 |
| At least once per week | 274 | 73.7 | 453 | 69.1 | 16 | 31.4 | 743 | 68.9 | 727 | 76.9 | 1,058 | 77.6 | 14 | 32.6 | 1,799 | 76.5 |
| Unknown | 44 | 11.8 | 81 | 12.3 | 26 | 51.0 | 151 | 14 | 72 | 7.6 | 117 | 8.6 | 22 | 51.2 | 211 | 9 |
| How often do you listen to the radio |  |  |  |  |  |  |  |  |  |  |  |  |  |  |  |  |
| Never | 45 | 12.1 | 79 | 12 | 6 | 11.8 | 130 | 12 | 55 | 5.8 | 113 | 8.3 | 6 | 14 | 174 | 7.4 |
| Less than weekly | 57 | 15.3 | 154 | 23.5 | 7 | 13.7 | 218 | 20.2 | 128 | 13.5 | 244 | 17.9 | 2 | 4.7 | 374 | 15.9 |
| At least once per week | 225 | 60.5 | 341 | 52 | 13 | 25.5 | 579 | 53.7 | 682 | 72.2 | 865 | 63.4 | 15 | 34.9 | 1,562 | 66.4 |
| Unknown | 45 | 12.1 | 82 | 12.5 | 25 | 49.0 | 152 | 14.1 | 80 | 8.5 | 142 | 10.4 | 20 | 46.5 | 242 | 10.3 |
| How often do you use the internet |  |  |  |  |  |  |  |  |  |  |  |  |  |  |  |  |
| Never | 3 | 0.8 | 2 | 0.3 | 2 | 3.9 | 7 | 0.6 | 2 | 0.2 | 5 | 0.4 | 1 | 2.3 | 8 | 0.3 |
| Less than weekly | 19 | 5.1 | 65 | 9.9 | 6 | 11.8 | 90 | 8.3 | 33 | 3.5 | 47 | 3.4 | 2 | 4.7 | 82 | 3.5 |
| At least once per week | 314 | 84.4 | 527 | 80.3 | 19 | 37.3 | 860 | 79.7 | 851 | 90.1 | 1,223 | 89.7 | 19 | 44.2 | 2,093 | 89 |
| Unknown | 36 | 9.7 | 62 | 9.5 | 24 | 47.1 | 122 | 11.3 | 59 | 6.2 | 89 | 6.5 | 21 | 48.8 | 169 | 7.2 |
| How often do you use social media platforms |  |  |  |  |  |  |  |  |  |  |  |  |  |  |  |  |
| Never | 3 | 0.8 | 11 | 1.7 | 3 | 5.9 | 17 | 1.6 | 6 | 0.6 | 2 | 0.1 | 1 | 2.3 | 9 | 0.4 |
| Less than weekly | 19 | 5.1 | 40 | 6.1 | 5 | 9.8 | 64 | 5.9 | 45 | 4.8 | 45 | 3.3 | 2 | 4.7 | 92 | 3.9 |
| At least once per week | 316 | 84.9 | 540 | 82.3 | 19 | 37.3 | 875 | 81.1 | 838 | 88.7 | 1,231 | 90.2 | 19 | 44.2 | 2,088 | 88.8 |
| Unknown | 34 | 9.1 | 65 | 9.9 | 24 | 47.1 | 123 | 11.4 | 56 | 5.9 | 86 | 6.3 | 21 | 48.8 | 163 | 6.9 |
| **Sexual experience** |  |  |  |  |  |  |  |  |  |  |  |  |  |  |  |  |
| **Relationship status** |  |  |  |  |  |  |  |  |  |  |  |  |  |  |  |  |
| Not in a relationship | 176 | 47.3 | 288 | 43.9 | 30 | 58.8 | 494 | 45.8 | 446 | 47.2 | 493 | 36.1 | 20 | 46.5 | 959 | 40.8 |
| In a relationship | 108 | 29 | 200 | 30.5 | 8 | 15.7 | 316 | 29.3 | 339 | 35.9 | 555 | 40.7 | 11 | 25.6 | 905 | 38.5 |
| Ever married/lived with someone | 4 | 1.1 | 4 | 0.6 | 9 | 17.6 | 17 | 1.6 | 24 | 2.5 | 51 | 3.7 | 4 | 9.3 | 79 | 3.4 |
| Unknown | 84 | 22.6 | 164 | 25 | 4 | 7.8 | 252 | 23.4 | 136 | 14.4 | 265 | 19.4 | 8 | 18.6 | 409 | 17.4 |
| **Ever had sex** |  |  |  |  |  |  |  |  |  |  |  |  |  |  |  |  |
| No | 157 | 42.2 | 258 | 39.3 | 12 | 23.5 | 427 | 39.6 | 422 | 44.7 | 440 | 32.3 | 2 | 4.7 | 864 | 36.7 |
| Yes | 110 | 29.6 | 208 | 31.7 | 4 | 7.8 | 322 | 29.8 | 368 | 38.9 | 627 | 46.0 | 13 | 30.2 | 1,008 | 42.9 |
| Prefer not to say | 22 | 5.9 | 27 | 4.1 | 33 | 64.7 | 82 | 7.6 | 26 | 2.8 | 41 | 3.0 | 23 | 53.5 | 90 | 3.8 |
| Unknown | 83 | 22.3 | 163 | 24.8 | 2 | 3.9 | 248 | 23.0 | 129 | 13.7 | 256 | 18.8 | 5 | 11.6 | 390 | 16.6 |

#### Reported sexual experience

Similarly, younger respondents were more likely to skip questions about sex and relationships: 31% of younger and 20% of older participants did not answer if they had ever had sex.(Table 2) Of those who responded, 43% of younger and 53% of older respondents reported ever having had sex, and 38% of younger and 47% of older respondents were currently in a relationship. A small minority (3%) had ever been married or co-habited with a partner. Among those who answered the questions, females were more likely than males to report ever having had sex (54% vs 45%), and being in a relationship (48% vs 41%). However, female respondents were more likely than males to skip questions about sex and relationships: 21% of female participants and 16% of males did not answer both questions.

#### Exposure to MTV Shuga Down South 2 and other MTV Shuga campaigns

Almost one-quarter (24%) of all respondents reported engagement with the campaign linked to the most recent MTV Shuga series: ‘Down South Season 2’ (DS2): 29% of the younger and 22% of older respondents.(Table 3) Engagement most often involved attending a peer educator-led group discussion about DS2 (15%), followed by watching the affiliated ‘In Real Life’ documentary (11%), reading the DS2 graphic novel (10%), attending a DS2 community event (9%), and watching DS2 on television (TV) or the internet and identifying it as season 2 (7%). Only a small proportion reported listening to DS2 on the radio (2%).

**Table 3.**
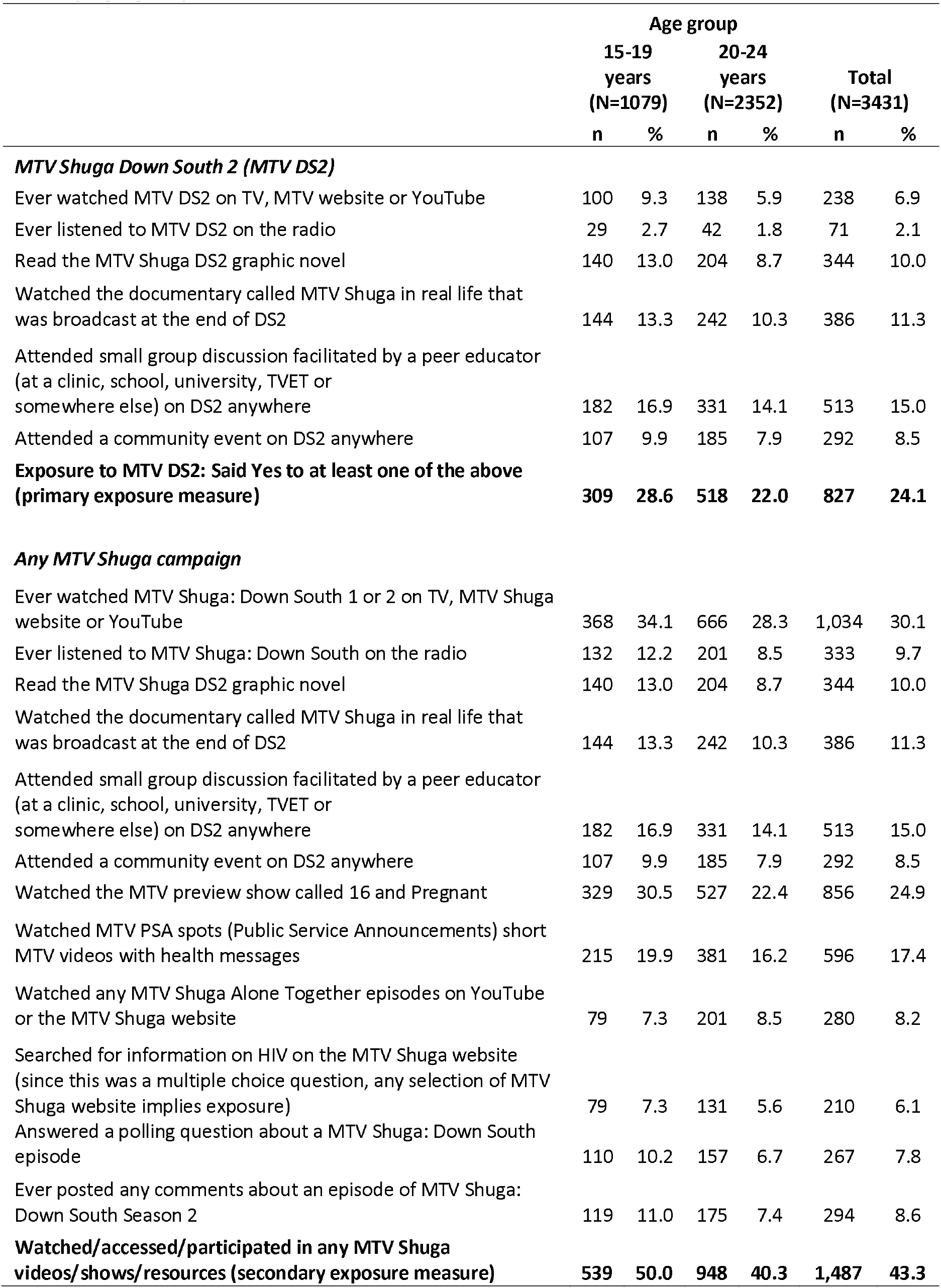
Proportions exposed to MTV Down South 2 and any MTV Shuga campaign, overall and by age group.

Overall, 43% (50% of the younger and 40% of the older respondents) had engaged with any MTV Shuga campaign.(Table 3) This included 30% of all respondents who ever watched ‘MTV Shuga Down South’ (either season 1 or 2) on TV or internet; 25% who watched the MTV preview show ‘16 and Pregnant’; 17% who watched MTV public service announcements related to Down South; as well as those who listened to either season 1 or 2 of Down South on the radio (10%), watched the COVID-19 related ‘Alone Together’ mini-series (8%), or interacted with the campaign by posting comments about an episode on social media (9%), answering a phone-based polling question (8%), or searching the MTV Shuga website (6%).

Engagement with MTV Shuga was higher among younger respondents (15-19 year-olds versus 20-24 year-olds) for the DS2 series specifically and any MTV Shuga campaign more broadly, and for all media formats and engagement activities.(Tables 4, S1) Exposure to MTV Shuga was also higher among females versus males, and to a lesser extent, those in school or university compared to those in technical/vocational colleges or out-of-school; and those who speak English or Zulu at home, compared to isiXhosa. Exposure was much higher among those who had ever been sexually active compared to those who had not, and for those currently in a relationship (for both DS2 and any MTV Shuga campaign).

**Table 4.** Factors associated with exposure to MTV Down South 2 and any MTV Shuga campaign.

|  | Exposure to Down South 2 |  |  |  |  |  | Exposure to any MTV Shuga campaign |  |  |  |  |  |
| --- | --- | --- | --- | --- | --- | --- | --- | --- | --- | --- | --- | --- |
|  | Total | Outcome | Row % | age-adjusted OR<sup>†</sup> |  |  | Total | Outcome | Row % | age-adjusted OR<sup>†</sup> |  |  |
|  | N | n | % | aOR | (95% CI) |  | N | n | % | aOR | (95% CI) |  |
| **Overall** | 3,198 | 805 | 25.2 |  |  |  | 3,198 | 1,458 | 45.6 |  |  |  |
| **Age group** |  |  |  |  |  |  |  |  |  |  |  |  |
| 15-19 | 988 | 300 | 30.4 | 1 |  |  | 988 | 527 | 53.3 | 1 |  |  |
| 20-24 | 2,210 | 505 | 22.9 | 0.68 | 0.57 | 0.80 | 2,210 | 931 | 42.1 | 0.64 | 0.55 | 0.74 |
| **Gender** |  |  |  |  |  |  |  |  |  |  |  |  |
| Male | 1,233 | 257 | 20.8 | 1 |  |  | 1,233 | 455 | 36.9 | 1 |  |  |
| Female | 1,874 | 538 | 28.7 | 1.50 | 1.27 | 1.78 | 1,874 | 984 | 52.5 | 1.86 | 1.61 | 2.16 |
| Other | 91 | 10 | 11.0 | 0.42 | 0.21 | 0.82 | 91 | 19 | 20.9 | 0.39 | 0.23 | 0.66 |
| **Current schooling/employment status** |  |  |  |  |  |  |  |  |  |  |  |  |
| In school (primary/secondary) | 653 | 223 | 34.2 | 1 |  |  | 653 | 386 | 59.1 |  |  |  |
| TVET<sup>‡</sup> | 921 | 120 | 13.0 | 0.31 | 0.23 | 0.42 | 921 | 196 | 21.3 | 0.2 | 0.16 | 0.26 |
| University | 1,094 | 369 | 33.7 | 1.06 | 0.83 | 1.35 | 1,094 | 646 | 59 | 1.08 | 0.85 | 1.36 |
| Other | 428 | 78 | 18.2 | 0.47 | 0.34 | 0.65 | 428 | 191 | 44.6 | 0.61 | 0.46 | 0.8 |
| Unknown | 102 | 15 | 14.7 | 0.35 | 0.2 | 0.62 | 102 | 39 | 38.2 | 0.45 | 0.29 | 0.69 |
| **Language spoken at home** |  |  |  |  |  |  |  |  |  |  |  |  |
| English | 231 | 74 | 32.0 | 1 |  |  | 231 | 133 | 57.6 | 1 |  |  |
| isiXhosa | 2,563 | 629 | 24.5 | 0.71 | 0.53 | 0.96 | 2,563 | 1,129 | 44.0 | 0.6 | 0.46 | 0.79 |
| Zulu | 216 | 65 | 30.1 | 0.89 | 0.59 | 1.33 | 216 | 116 | 53.7 | 0.83 | 0.57 | 1.21 |
| Other<sup>§</sup> | 188 | 37 | 19.7 | 0.52 | 0.33 | 0.81 | 188 | 80 | 42.6 | 0.54 | 0.36 | 0.8 |
| **Urban/Rural residence** |  |  |  |  |  |  |  |  |  |  |  |  |
| Urban setting | 2,741 | 686 | 25.0 | 1 |  |  | 2,741 | 1,232 | 44.9 | 1 |  |  |
| Rural setting | 255 | 81 | 31.8 | 1.37 | 1.04 | 1.81 | 255 | 152 | 59.6 | 1.78 | 1.37 | 2.32 |
| Unknown | 202 | 38 | 18.8 | 0.66 | 0.45 | 0.94 | 202 | 74 | 36.6 | 0.66 | 0.49 | 0.89 |
| **Province** |  |  |  |  |  |  |  |  |  |  |  |  |
| Eastern Cape (EC) - Mthatha | 2,305 | 573 | 24.9 | 1 |  |  | 2,305 | 969 | 42.0 | 1 |  |  |
| Eastern Cape - O.R. Tambo or other EC | 321 | 94 | 29.3 | 1.22 | 0.94 | 1.58 | 321 | 189 | 58.9 | 1.93 | 1.52 | 2.45 |
| Other provinces | 509 | 120 | 23.6 | 0.89 | 0.71 | 1.12 | 509 | 265 | 52.1 | 1.43 | 1.18 | 1.74 |
| Unknown | 63 | 18 | 28.6 | 1.18 | 0.68 | 2.06 | 63 | 35 | 55.6 | 1.68 | 1.02 | 2.79 |
| **Food insecurity** |  |  |  |  |  |  |  |  |  |  |  |  |
| Never/rarely | 1,785 | 371 | 20.8 | 1 |  |  | 1,785 | 735 | 41.2 | 1 |  |  |
| Sometimes | 1,048 | 321 | 30.6 | 1.71 | 1.44 | 2.04 | 1,048 | 550 | 52.5 | 1.61 | 1.38 | 1.88 |
| Often/always | 204 | 77 | 37.7 | 2.31 | 1.70 | 3.14 | 204 | 121 | 59.3 | 2.09 | 1.55 | 2.81 |
| Unknown | 161 | 36 | 22.4 | 1.05 | 0.71 | 1.56 | 161 | 52 | 32.3 | 0.65 | 0.46 | 0.91 |
| **Household media assets index** |  |  |  |  |  |  |  |  |  |  |  |  |
| Low | 1,176 | 320 | 27.2 | 1 |  |  | 1,176 | 570 | 48.5 | 1 |  |  |
| Medium | 1,048 | 217 | 20.7 | 0.72 | 0.59 | 0.88 | 1,048 | 439 | 41.9 | 0.79 | 0.67 | 0.94 |
| High | 974 | 268 | 27.5 | 1.03 | 0.85 | 1.25 | 974 | 449 | 46.1 | 0.92 | 0.78 | 1.09 |
| **Individual media assets index** |  |  |  |  |  |  |  |  |  |  |  |  |
| Low | 1,190 | 250 | 21.0 | 1 |  |  | 1,190 | 503 | 42.3 | 1 |  |  |
| Medium | 1,218 | 341 | 28.0 | 1.50 | 1.24 | 1.81 | 1,218 | 638 | 52.4 | 1.54 | 1.31 | 1.82 |
| High | 790 | 214 | 27.1 | 1.47 | 1.19 | 1.82 | 790 | 317 | 40.1 | 0.96 | 0.80 | 1.16 |
| **Relationship status** |  |  |  |  |  |  |  |  |  |  |  |  |
| Not in a relationship | 1,453 | 287 | 19.8 | 1 |  |  | 1,453 | 528 | 36.3 | 1 |  |  |
| In a relationship | 1,221 | 313 | 25.6 | 1.45 | 1.21 | 1.74 | 1,221 | 604 | 49.5 | 1.80 | 1.54 | 2.10 |
| Ever married/lived with someone | 96 | 24 | 25.0 | 1.45 | 0.90 | 2.35 | 96 | 45 | 46.9 | 1.68 | 1.11 | 2.56 |
| Unknown | 428 | 181 | 42.3 | 2.96 | 2.35 | 3.73 | 428 | 281 | 65.7 | 3.34 | 2.66 | 4.20 |
| **Ever had sex** |  |  |  |  |  |  |  |  |  |  |  |  |
| No | 1,291 | 162 | 12.5 | 1 |  |  | 1,291 | 286 | 22.2 | 1 |  |  |
| Yes | 1,330 | 453 | 34.1 | 3.84 | 3.13 | 4.70 | 1,330 | 868 | 65.3 | 7.52 | 6.28 | 9.01 |
| Prefer not to say | 172 | 12 | 7.0 | 0.48 | 0.26 | 0.89 | 172 | 27 | 15.7 | 0.57 | 0.37 | 0.89 |
| Unknown | 405 | 178 | 44.0 | 5.41 | 4.18 | 7.00 | 405 | 277 | 68.4 | 7.78 | 6.05 | 10.01 |
| **Called a helpline or searched for information on HIV on the internet** |  |  |  |  |  |  |  |  |  |  |  |  |
| No | 1,667 | 296 | 17.8 | 1 |  |  | 1,667 | 495 | 29.7 | 1 | 1 | 1 |
| Yes | 1,352 | 456 | 33.7 | 2.35 | 1.98 | 2.78 | 1,352 | 888 | 65.7 | 4.57 | 3.91 | 5.33 |
| Unknown | 179 | 53 | 29.6 | 1.84 | 1.30 | 2.60 | 179 | 75 | 41.9 | 1.59 | 1.15 | 2.18 |

<sup>†</sup>OR = Odds Ratio; <sup>‡</sup>TVET - Technical and Vocational Education and Training; <sup>§</sup>includes 5 respondents whose language was unknown

#### HIV outcomes and associations with MTV Shuga exposure

About 40% of respondents said they were aware of their HIV status, either because they had ever tested HIV-positive (∼11%; 136 out of 1294 who knew their status) or because they had tested in the past year and received their result.(Table 5) Knowledge of HIV status was higher among those who were exposed to MTV Shuga DS2 (58%) compared to those who were not (35%) (adjusted odds ratio (aOR)=2.06 [95% confidence interval 1.64-2.58]), after controlling for possible confounders, including alternative sources of HIV information.(Table 5, Figure 1a, Table S2) The association between DS2 exposure and knowledge of HIV status was stronger among older respondents (aOR=2.61) than younger respondents (aOR=1.60; p<0.001 for test of interaction by age group).

**Table 5.**
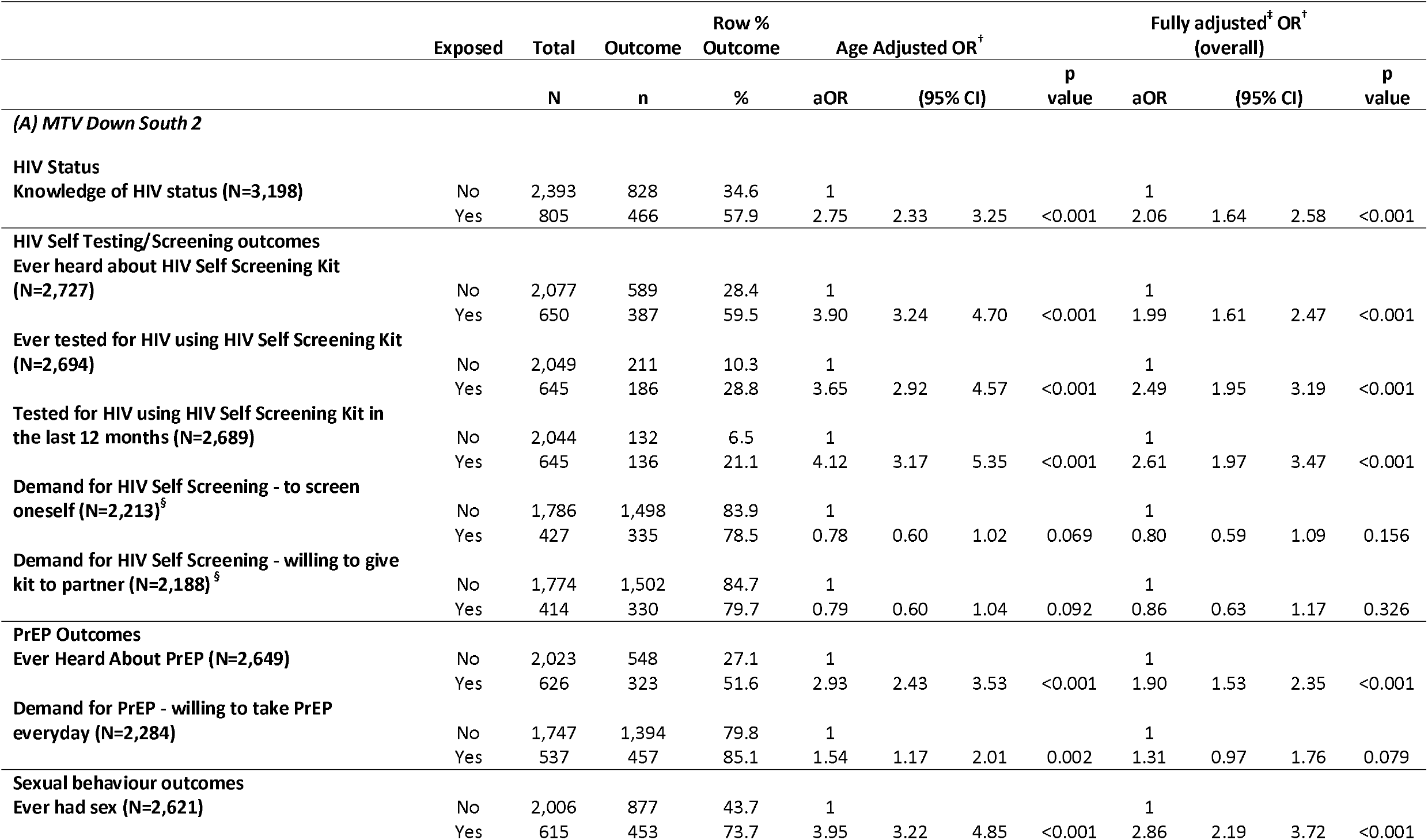
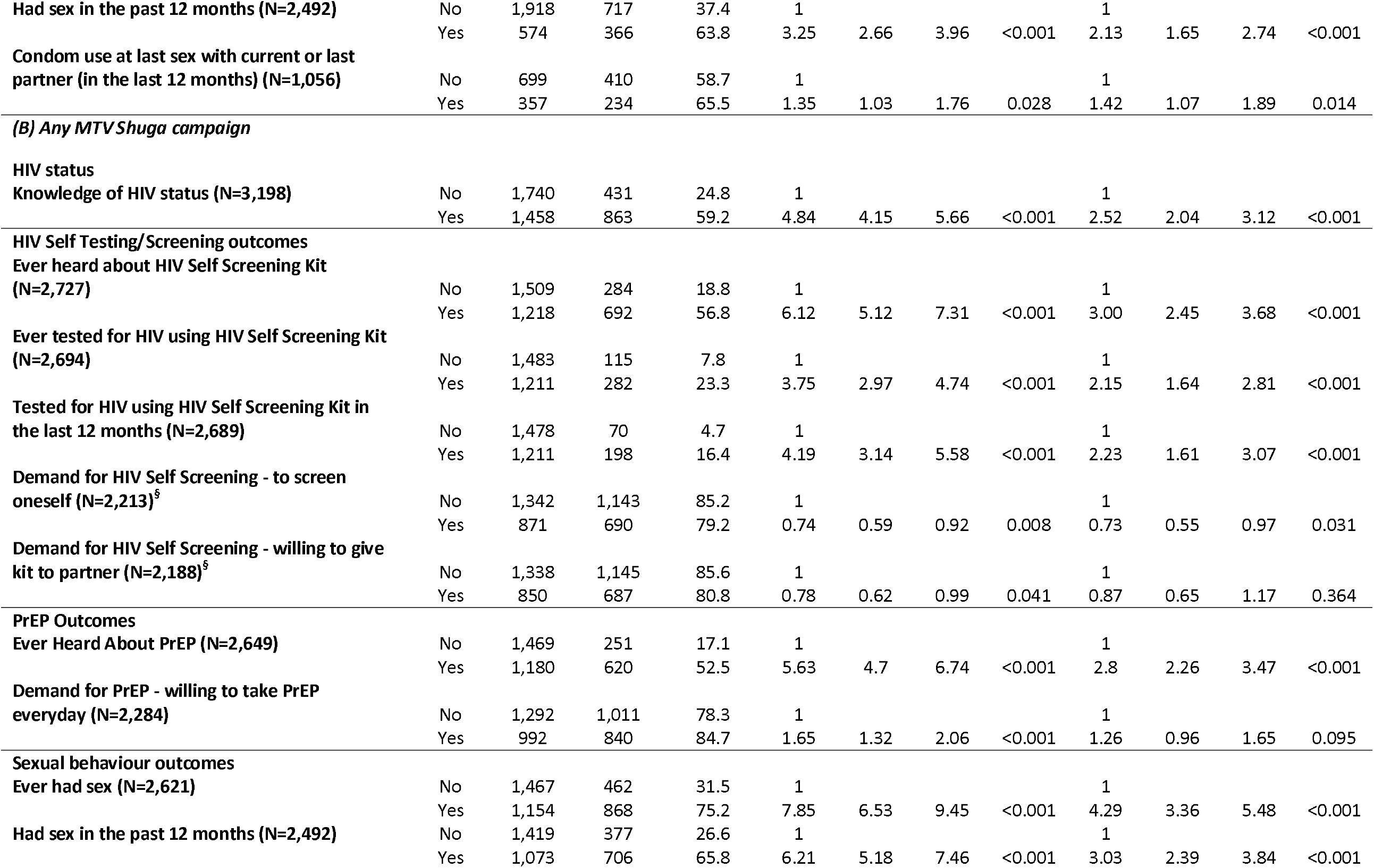
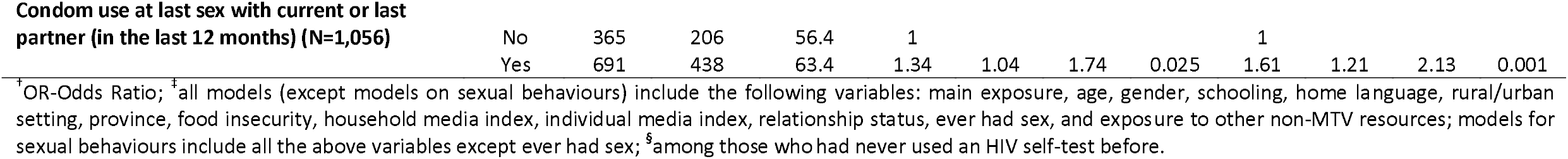
Associations between exposure to MTV Down South 2 (A) and any MTV Shuga campaign (B) with HIV self-testing and PrEP outcomes.

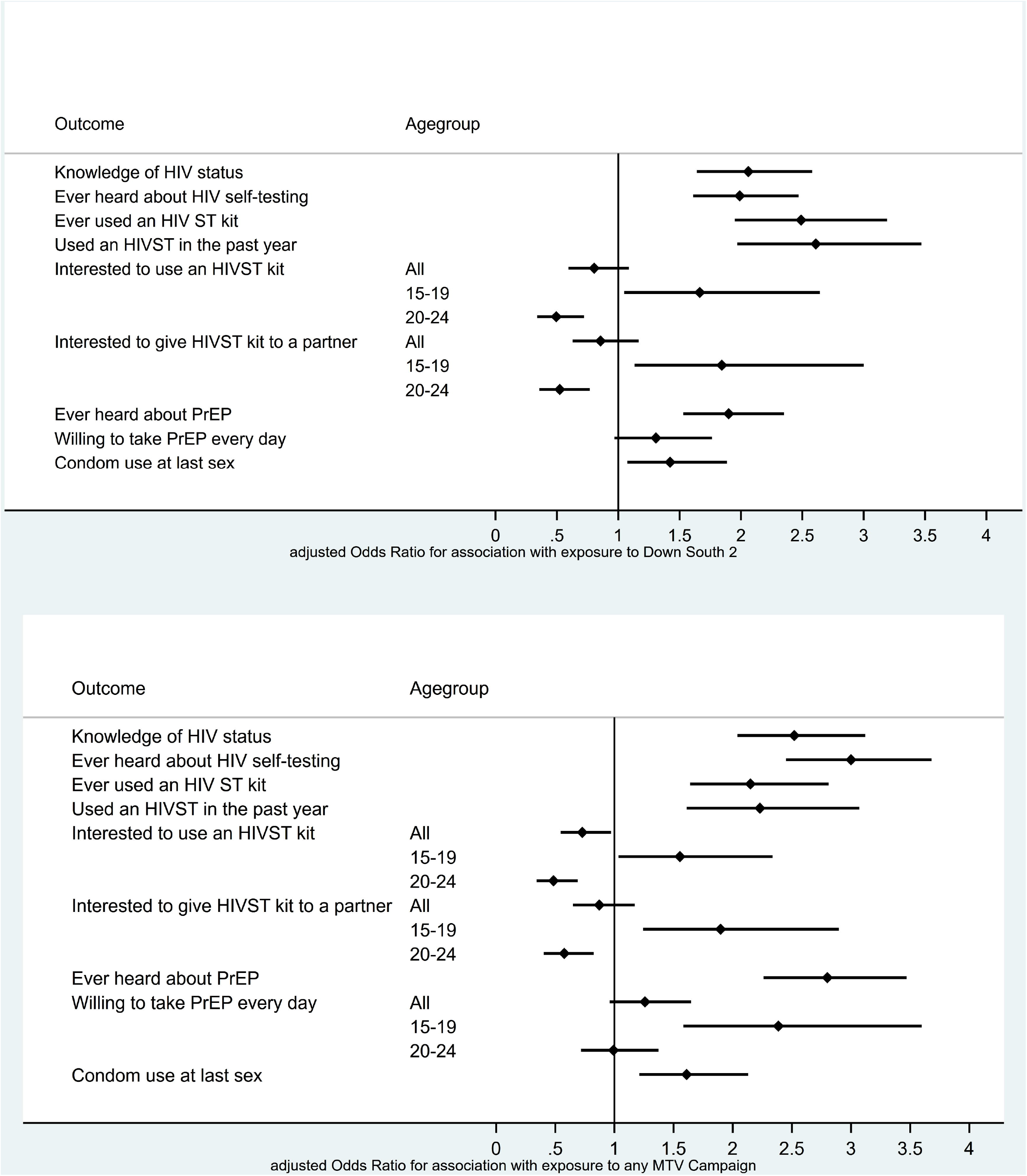

The proportion of respondents who had ever heard of HIV self-testing was also higher among those exposed to DS2 versus the non-exposed (60% vs 28%; aOR=1.99 [1.61-2.47]), with a stronger effect size among older versus younger respondents (p<0.001).(Table 5, Figure 1a) The proportions who had used an HIV self-test, either ever in their lifetime (29% vs 10%; aOR=2.49 [1.95-3.19]) or within the past year (21% vs 7%; aOR=2.61 [1.97-3.47]) were higher among those exposed versus non-exposed to DS2, with no evidence of a difference by age group. Among those who had never used an HIV self-test before, interest in using a self-test was high overall (83%), and association with DS2 exposure differed by age group: specifically, exposure to DS2 was associated with more interest in HIV self-testing among younger respondents (aOR=1.66 [1.05-2.64]) and with less interest among older respondents (aOR=0.50 [0.34-0.72]). Similarly, interest in giving an HIV self-test kit to a partner was high, with DS2 exposure associated with more interest among younger respondents (aOR=1.84 [1.13-3.0]) and with less interest among the older (aOR=0.52 [0.36-0.77]), compared to the non-exposed.

One-third of respondents were aware of PrEP, with higher proportions aware of PrEP among those exposed versus not exposed to DS2 (52% vs 27%; aOR=1.90 [1.53-2.35]). Demand for PrEP (i.e., willingness to take PrEP every day) was high overall with weak evidence of higher demand among those exposed to DS2 (85% vs 80% among non-exposed; aOR=1.31 [0.97-1.76]), and no differences by age group.(Table 5, Table S2)

Respondents who were exposed to DS2 were more likely to have had sex, ever in their lifetime (74% vs 44% of the non-exposed; aOR=2.86 [2.19-3.72]) and in the past 12 months (64% vs 37%); aOR=2.13 [1.65-2.74]), than non-exposed respondents. This was particularly true for older respondents, with those exposed to DS2 much more likely to have ever had sex than those not exposed (aOR=8.05 [5.57-11.65]). Among those who reported sex within the past 12 months, DS2 audiences were more likely to report condom use at last sex than those not exposed to DS2 (66% vs 59%; aOR=1.42 [1.07-1.89]).(Table 5, Table S2)

In sensitivity analyses in which multiple imputation was conducted for all participants with missing values, results were consistent with the complete case analysis.(Table S3) In analysis of the secondary exposure measure (exposure to any MTV Shuga campaign, including but not limited to DS2), most outcomes were more common among those exposed to any MTV Shuga compared to those with no MTV Shuga exposure of any kind, and associations between exposure and outcomes were similar to those observed with exposure to MTV Shuga DS2 specifically.(Table 5B, Figure 1b)

In more detailed reports of HIV testing experiences, those exposed to DS2 accessed HIV testing services more frequently than those not exposed (ever and within the past year). (Table S4) This was true of HIV self-testing as well as testing with a community health worker and within ante-natal care services (for females). HIV self-test kits were accessed from a range of outlets, most often government clinics, community health centres, or hospitals, and university/TVET settings. Pharmacies were also a common source, particularly for younger (15-19 year-old) participants, and more so among young DS2 audiences (27%) than those not exposed (17%). DS2 viewers were also more likely to have paid fees for their last self-test (36% vs 21% of non-exposed), among both age groups. Respondents cited a range of reasons for their last HIV self-test, with the most common motivation being to learn their HIV status (>80% of those who self-tested). Of those exposed to DS2, 32% of the younger respondents and 18% of older respondents cited MTV Shuga as the reason they got their most recent HIV self-test. Following their self-test, DS2 viewers were more likely than non-viewers to seek a laboratory test to confirm the result (66% vs 56% of self-testers).

### Qualitative research

In analysis of qualitative data, themes emerged about the behavioural mechanisms through which MTV Shuga DS2 influenced participants’ capability, motivation and opportunity related to HIV self-testing and PrEP outcomes.(Table S5) **Awareness** (an aspect of capability) of HIVST and PrEP was enhanced by watching the show, with some learning about these resources for the first time from DS2 –*”Honestly speaking, I didn’t know about the self-testing until I watched the show now. I now know that you can test yourself”*– while others gained a greater understanding through the show: *“Before seeing the show, PrEP was just something I saw in books. I didn’t know that it is actually out there*.*”* Participants expressed ways in which this understanding, and storylines (about Bongi and Arrabella living with HIV), helped to reduce their fear about HIV and knowing their own status, *“The show showed me that knowing your status is better than just living with an unknown status… [it] won’t mean the end of the world*.*”* Some also gained **confidence** to enquire and learn more, e.g., from internet searches and approaching health service staff. Young people described how scenes and storylines from DS2 helped them feel more **prepared** to access HIV services: *“I know what I will be experiencing from the whole situation*.*”* This included participants who were not yet sexually active and felt better prepared for future relationships, *“Now that I have information about PrEP, it will help me when I meet someone. I will know what I need to do and know how PrEP will help me*.*”* Scenarios presented through the show also helped young people to **reflect** on their own needs and preferences for HIV prevention options, at different times of their life. After watching the show, some realised they prefer self-testing at home, for the privacy and convenience, while others would be more anxious testing at home and would prefer the support available at a clinic: *“For me, this thing of testing at home really scared me… At home, there will be no one to help you [after getting the results]*.*”* Participants were also able to imagine scenarios in which they could use PrEP, e.g., *“I like the fact that it protects you from getting HIV when your partner has tested positive,”* though some preferred to rely on condoms for HIV prevention. While capability and motivation were clearly enhanced for many, the show had less influence on **opportunity to access** services like HIVST and PrEP. While they saw ways in which they could get such resources, from the show, many doubted the availability in their own setting *(“I wouldn’t be confident that if I go to the hospital, I’d get them. The hospitals here are not the same as those in Joburg*.*”*), especially during closures related to COVID-19 lockdowns.

The qualitative data also illustrated ways in which DS2 had a broader influence, beyond its immediate audience.(Table S6) It helped some viewers initiate conversations and discussion with partners and parents, with whom they said conversations about sex are often avoided or awkward, although some remained hesitant to engage in such discussions. Some young males described ways in which the show changed the tone of conversations with peers, to more substantive discussions about PrEP and contraception. Many felt enlightened by the show and wished more young people would have access to the information about PrEP and HIV self-screening. Some felt that this should be done through more government and health campaigns, and many saw the role they themselves could play as educators and advocates for HIV testing and prevention: *“People are usually afraid of the pain and pricking when it comes to taking HIV test. So, I’ll definitely be an advocate for the self-screening HIV test now because of the show,”* and within their own family: *“I can be able to give my little brother advice on when he wants to date*… *Now I can explain to him how to be safe when he’s having sex, testing, everything*.*”*

Some participants said they were unaffected by the show and didn’t learn anything about PrEP and HIV self-testing. These topics featured in four different scenes (8 minutes of content) in the 10 episodes of the DS2 series, and some viewers missed those scenes, or were distracted at the time. Some were confused by the scenes and didn’t gain confidence or motivation to use PrEP or self-testing: *“Since they say HIV is something that is tested through blood, I kind of don’t understand what they are doing there [with HIV self-screening] ‘cause there’s only saliva inside the mouth*.*”*

## Discussion

Through a mixed methods evaluation of MTV Shuga’s multimedia edutainment campaign, we found evidence consistent with a causal impact of the campaign on important HIV prevention outcomes among young people in South Africa. Among 3,431 15-24 year-old participants of an online survey, based predominantly in Eastern Cape province, substantial proportions had been exposed to MTV Shuga: one-quarter had engaged with the most recent MTV Shuga series (Down South 2; DS2) and 43% with any component or past series of the MTV Shuga campaign. Exposure to DS2 and the wider MTV Shuga campaign was strongly associated with the primary outcome of the study, knowledge of HIV status, as well as increased awareness and use of HIV self-testing (HIVST) and knowledge of PrEP. Respondents’ interest to use HIVST and PrEP was very high overall; it was higher among MTV Shuga audiences for the younger (15-19 year-olds) but not among older respondents aged 20-24 years.

An embedded qualitative study offered insights into ways in which MTV Shuga DS2 may have influenced these outcomes. DS2 enhanced young people’s capability and motivation to adopt HIV prevention behaviours, while its influence on opportunity (e.g., to access prevention services) was more limited.[20] Some viewers were introduced to HIV self-testing and PrEP for the first time through the show, and greater awareness helped to reduce fear and boost confidence to learn their HIV status and seek further HIV information and services. For others, HIV information that had previously seemed abstract or academic was made real and relevant in DS2 scenarios and storylines, with viewers given an opportunity to observe behaviours that were typically kept private, e.g., couples self-testing together at home. This was evidence of observational learning, consistent with Social Learning Theory.[22] While not all felt they currently needed to use resources like HIV self-testing and PrEP, they imagined future circumstances in which they would benefit from them. Some reflected on options presented by DS2 storylines, to think through their personal preferences for HIV prevention, and this may help to explain the differences in demand for HIV self-testing by age group, e.g., if older viewers generally valuing the support available from provider-led testing and younger adolescents preferring the privacy of self-testing at home. For some, the scenes on HIVST and PrEP were too short or unclear to have any influence.

The findings suggest ways in which a popular, immersive multimedia campaign like MTV Shuga can accelerate achievement of HIV prevention goals for young people. MTV Shuga’s influence is consistent with a person-centred approach to HIV prevention: through engaging storylines and characters, young people are offered accurate and relevant information on HIV prevention options and enabled to make their own choices (in contrast to assuming that young people are ‘passive beneficiaries’).[23] Respecting personal choice and agency in this way can help people to adopt HIV prevention choices at different time points, as their needs and situations change, analogous to offering a ‘contraceptive method mix’ reflecting users’ diverse preferences and circumstances.[24] This can help young people to prepare for changes that may increase their HIV risk, for example, to know prevention options before their first sexual experience. In southern Africa, many young people do not engage with SRH services until they are already sexually active, pregnant or married, at which point many are already at risk for HIV/STI (in settings where HIV incidence escalates from an early age) [25, 26]. The first sexual experience is often one in which young people are not well prepared, e.g., with information, contraception, condoms or HIV/STI testing.[27] While MTV Shuga audiences were more likely to have had sex than those who never engaged with the campaign, viewers who had never had sex felt more aware and prepared for HIV prevention choices once they became sexually active. Acquiring a new sexual partner is another high-risk transition point for young people, for which they can be prepared in advance, and MTV Shuga viewers anticipated prevention choices they could make with new partners, e.g., to use PrEP with a partner who is living with HIV.

The benefits of MTV Shuga may be greatest for those who are most connected to digital media for access to the dramatic television series with high production values, and the ability to be immersed in all episodes. However, in the study setting of Mthatha, more respondents reported experiencing ‘offline’ components of the DS2 campaign than the televised or internet-streamed episodes. Many had engaged with community-based viewing events, peer-led Shuga discussions, and the DS2 graphic novels distributed through schools. Our qualitative research found that Shuga opened up dialogue about sex and HIV with peers, parents and partners, and this ability for Shuga to stimulate real-life discussions can happen across all its formats. Also, some viewers were inspired to become ‘advocates’ for the new HIV prevention information they learned via DS2, suggesting that, by reaching the most digitally connected young people, MTV Shuga can inspire early adopters of HIV preventive innovations, who are well placed to diffuse those messages to peers online and in real life.[28] An important limitation to a multimedia campaign like MTV Shuga is its lack of influence on supply of services, either actual or perceived supply, including the HIV prevention technologies it promoted through DS2. Viewers were typically sceptical about the opportunity to access HIV self-testing and PrEP in their area of Eastern Cape, despite recent investments in HIV self-test and PrEP distribution.[18] The survey showed that DS2 viewers were more likely than non-viewers to have paid a fee for their last HIV self-test kit, suggesting that the ability to pay or access a pharmacy may help to explain the observed differences in use of self-testing, even if MTV Shuga boosted motivation for HIV testing. To ensure equitable access to prevention tools and services, media campaigns that generate demand should be directly linked to distribution efforts that are available and acceptable to young people.

The findings are consistent with a small but growing number of studies that demonstrate benefits of MTV Shuga. The afore-mentioned trial in Nigeria found increases in HIV and STI testing in communities randomly selected to receive group showings of MTV Shuga episodes.[15] Among a cohort of adolescent girls and young women in rural KwaZulu, exposure to MTV Shuga was relatively low by 2019 (∼15% of randomly sampled 15-24 year old females), but associated with a range of sexual and reproductive health benefits (although not with HIV testing, perhaps due to regular HIV surveillance in the study area).[17] In this study, the higher prevalence of exposure to MTV Shuga (>40%) may be due to better media access as well as more concerted efforts to offer MTV Shuga activities offline, through schools and communities in Eastern Cape.

Our study had a number of limitations. It is likely that the sample does not represent the general population of Eastern Cape, but is skewed in favour of those young people who frequently engage with digital media. Compared to a representative sample in urban areas of Eastern Cape in 2016, cell-phone ownership was similar among 20-24 year olds, but higher among our younger respondents (15-19 years).[29] This may reflect recent growth in phone ownership among teenagers, throughout South Africa and Mthatha. Only 1% of our study participants reported that they never used the internet. This may reflect the virtual, internet-based study methods we used to comply with COVID-19 restrictions in South Africa and avoid SARS-CoV-2 transmission risk, however, connectedness and social media use is rapidly growing across South Africa. A recent study in Mthatha described young people’s social media use as “pervasive”.[30] We tried to minimise cost barriers to participation, by making the internet survey free to access (through reverse-charging) and providing data credit to all participants, however, participation was most likely easier for those with digital devices, more reliable internet access and greater literacy.

Our design also relied on self-reported measures of MTV Shuga engagement and misclassification may have under- or over-estimated exposure. Many respondents opted not to answer some questions, particularly the questions on sexual experiences, suggesting that online surveys are not ideal for collecting such private information from young people, even if anonymised. However, the study findings were largely unchanged in sensitivity analyses with imputation of missing values. Another limitation is that the increased associations observed may be explained by unknown or unmeasured confounders, especially as the questionnaire length was kept as short as possible for online use. Nevertheless, associations remained strong after accounting for numerous potential confounders, including other sources of HIV prevention information and campaigns, and were consistent across multiple outcomes. Also, the strengths of associations observed across outcomes, and the connections between mechanisms (identified in qualitative analysis) and the observed outcomes offered evidence of causality consistent with a realist evaluation.[31]

## Conclusions

As new HIV testing and PrEP options become available, we sought evidence of a multimedia edutainment campaign’s influence on awareness and demand for such tools, among young men and women in a high-prevalence setting of South Africa. Together, findings from a large online survey and embedded qualitative research are consistent with a causal impact of MTV Shuga on young people’s awareness and use of HIV self-testing and PrEP, and knowledge of their own HIV status, although we cannot exclude other possible explanations for the observed associations. With online and offline components, the ‘360-degree’ media campaign reached sizeable proportions of young people through TV, internet, schools and community events, especially adolescent girls – a group at particularly high risk of HIV acquisition in high-burden countries. Positive effects were equally strong for females and males, and the diverse characters and storylines enabled MTV Shuga viewers to reflect on their personal needs and HIV prevention choices. With stronger, more direct links to the supply and distribution of prevention tools like self-test kits and PrEP, growing use of digital media among young people, and the potential for early adopters to diffuse innovation, a popular edutainment campaign like MTV Shuga has an important role to play in closing age and gender gaps in HIV testing and prevention goals.

## Data Availability

All data produced in the present study are available upon reasonable request to the authors

## Additional information

### Authors’ contributions

IB, SC, SS, CC, DK, VB co-designed the study protocol, and CC, DK, DOD and VB managed the data collection. SS led the survey questionnaire development and DOD programmed the questionnaire. SM led the management and analysis of survey data, VB led the analysis of qualitative data, and IB led the triangulation. IB wrote the first draft, all authors contributed to revisions and reviewed the final draft.

## Acknowledgements

The authors wish to thank all the young people who contributed their time to this study. We thank Michel Carael, Oona Campbell, Hasina Subedar, Sian Floyd, Heather Ingold, Ombeni Mwerindi, Rachel Baggaley, Kathering Fielding and Melissa Newman for valuable ideas to strengthen this research. We are grateful to Antonio Duran-Aparicio for all the support to make this project possible. We acknowledge the dedication of MTV Shuga peer educators and coordinators (Yvonne Diogo, Lesedi Thwala) in South Africa and appreciate the trust of the MTV Staying Alive Foundation, especially Georgia Arnold and Sara Piot.

## Additional files

Figure S1. Activities in MTV Shuga Down South 2 360-degree media campaign

Figure S2. Hypothesised common causes of MTV Shuga exposure and HIV outcomes

Table S1. Factors associated with exposure to MTV Down South 2 and any MTV Shuga campaign, among 15-19 year-olds (A) and 20-24 year-olds (B)

Table S2. Associations between exposure to MTV Down South 2 (A) and any MTV Shuga campaign (B) with HIV self-testing and PrEP outcomes: Age specific estimates

Table S3. Results from multiple imputation with primary exposure: m=10 imputations

Table S4. Experiences with HIV self-testing and PrEP, among those ever and never exposed to MTV Shuga DS2

Table S5. Mechanisms by which MTV Shuga Down South 2 influenced behaviour through Capability, Motivation and Opportunity to improve HIV self-testing and PrEP outcomes: themes emerging from deductive analysis of qualitative research in Mthatha, Eastern Cape

Table S6. Evidence of broader social mechanisms by which MTV Shuga DS2 influenced HIV self-testing and PrEP outcomes: themes emerging from open code analysis of qualitative research in Mthatha, Eastern Cape

File S1. Online survey questionnaire

File S2. Topic guides for the qualitative research activities

