## Supplemental Tables 1-4 for "Effects of a multimedia campaign on HIV self-testing and PrEP outcomes among young people in South Africa: A mixed-methods impact evaluation of ‘MTV Shuga Down South’"

**Table S1: Factors associated with exposure to MTV Down South 2 and any MTV Sh**
**A: 15-19 years**

| | | | | Exposure to Down | |
|---|---|---|---|---|---|
| | Total | Outcome | Row % Outcome | Age Adjusted ( | |
| | N | n | % | aOR | (95 |
| **Overall** | 988 | 300 | 30.4 | | |
| **Gender** | | | | | |
| Male | 337 | 82 | 24.3 | 1 | |
| Female | 601 | 213 | 35.4 | 1.71 | 1.27 |
| Other | 50 | 5 | 10 | 0.35 | 0.13 |
| **Current schooling/employment status** | | | | | |
| In school (primary/secondary) | 536 | 193 | 36 | 1 | |
| TVET[§] | 137 | 22 | 16.1 | 0.34 | 0.21 |
| University | 214 | 71 | 33.2 | 0.88 | 0.63 |
| Other | 55 | 6 | 10.9 | 0.22 | 0.09 |
| Unknown | 46 | 8 | 17.4 | 0.37 | 0.17 |
| **Language spoken at home** | | | | | |
| English | 88 | 27 | 30.7 | 1 | |
| isiXhosa | 729 | 228 | 31.3 | 1.03 | 0.64 |
| Zulu | 97 | 31 | 32 | 1.06 | 0.57 |
| Other[¶] | 74 | 14 | 18.9 | 0.53 | 0.25 |
| **Urban/Rural residence** | | | | | |
| Urban setting | 814 | 265 | 32.6 | 1 | |
| Rural setting | 87 | 25 | 28.7 | 0.84 | 0.51 |
| Unknown | 87 | 10 | 11.5 | 0.27 | 0.14 |
| **Province** | | | | | |
| Eastern Cape (EC) - Mthatha | 650 | 217 | 33.4 | 1 | |
| Eastern Cape - OR Tambo or other EC | 113 | 31 | 27.4 | 0.75 | 0.48 |
| Other provinces | 203 | 46 | 22.7 | 0.58 | 0.41 |
| Unknown | 22 | 6 | 27.3 | 0.75 | 0.29 |
| **Food insecurity** | | | | | |
| Never/rarely | 561 | 158 | 28.2 | 1 | |
| Sometimes | 294 | 91 | 31 | 1.14 | 0.84 |
| Often/always | 67 | 34 | 50.7 | 2.63 | 1.57 |
| Unknown | 66 | 17 | 25.8 | 0.88 | 0.49 |
| **Household media assets index** | | | | | |
| Low | 406 | 117 | 28.8 | 1 | |
| Medium | 280 | 74 | 26.4 | 0.89 | 0.63 |
| High | 302 | 109 | 36.1 | 1.4 | 1.01 |

| | | | | | |
|---|---|---|---|---|---|
| **Individual media assets index** | | | | | |
| Low | 422 | 110 | 26.1 | 1 | |
| Medium | 373 | 117 | 31.4 | 1.3 | 0.95 |
| High | 193 | 73 | 37.8 | 1.73 | 1.2 |
| **Relationship status** | | | | | |
| Not in a relationship | 494 | 128 | 25.9 | 1 | |
| In a relationship/ever married/lived w so | 333 | 104 | 31.2 | 1.3 | 0.96 |
| Unknown | 161 | 68 | 42.2 | 2.09 | 1.44 |
| **Ever had sex** | | | | | |
| No | 427 | 114 | 26.7 | 1 | |
| Yes | 322 | 113 | 35.1 | 1.48 | 1.08 |
| Prefer not to say/unknown | 239 | 73 | 30.5 | 1.21 | 0.85 |
| **Called a helpline or searched for information on HIV on the internet** | | | | | |
| No | 485 | 125 | 16 | 1 | |
| Yes | 422 | 157 | 43.3 | 1.71 | 1.29 |
| Unknown | 81 | 18 | 22.2 | 0.82 | 0.47 |

†age-group specific Odds Ratios (OR); ‡only variables associated with the outcome at p<=0.1 were

**B: 20-24 years**

| | Exposure to Down South 2 | | | | |
|---|---|---|---|---|---|
| | Total | Outcome | Row % Outcome | Age Adjusted ( | |
| | N | n | % | aOR | (95% |
| **Overall** | 2,210 | 505 | 22.9 | | |
| **Gender** | | | | | |
| Male | 896 | 175 | 19.5 | 1 | |
| Female | 1,273 | 325 | 25.5 | 1.41 | 1.15 |
| Other | 41 | 5 | 12.2 | 0.57 | 0.22 |
| **Current schooling/employment status** | | | | | |
| In school (primary/secondary) | 117 | 30 | 25.6 | 1 | |
| TVET§ | 784 | 98 | 12.5 | 0.41 | 0.26 |
| University | 880 | 298 | 33.9 | 1.48 | 0.96 |
| Other | 373 | 72 | 19.3 | 0.69 | 0.43 |
| Unknown | 56 | 7 | 12.5 | 0.41 | 0.17 |
| **Language spoken at home** | | | | | |
| English | 143 | 47 | 32.9 | 1 | |
| isiXhosa | 1,834 | 401 | 21.9 | 0.57 | 0.4 |
| Zulu | 119 | 34 | 28.6 | 0.82 | 0.48 |
| Other¶ | 114 | 23 | 20.2 | 0.52 | 0.29 |

| | | | | | |
|---|---|---|---|---|---|
| **Urban/Rural residence** | | | | | |
| Urban setting | 1,927 | 421 | 21.8 | 1 | |
| Rural setting | 168 | 56 | 33.3 | 1.79 | 1.28 |
| Unknown | 115 | 28 | 24.3 | 1.15 | 0.74 |
| **Province** | | | | | |
| Eastern Cape (EC) - Mthatha | 1,655 | 356 | 21.5 | 1 | |
| Eastern Cape - OR Tambo or other EC | 208 | 63 | 30.3 | 1.59 | 1.15 |
| Other provinces | 306 | 74 | 24.2 | 1.16 | 0.87 |
| Unknown | 41 | 12 | 29.3 | 1.51 | 0.76 |
| **Food insecurity** | | | | | |
| Never/rarely | 1,224 | 213 | 17.4 | 1 | |
| Sometimes | 754 | 230 | 30.5 | 2.08 | 1.68 |
| Often/always | 137 | 43 | 31.4 | 2.17 | 1.47 |
| Unknown | 95 | 19 | 20.0 | 1.19 | 0.70 |
| **Household media assets index** | | | | | |
| Low | 770 | 203 | 26.4 | 1 | |
| Medium | 768 | 143 | 18.6 | 0.64 | 0.5 |
| High | 672 | 159 | 23.7 | 0.87 | 0.68 |
| **Individual media assets index** | | | | | |
| Low | 768 | 140 | 18.2 | 1 | |
| Medium | 845 | 224 | 26.5 | 1.62 | 1.27 |
| High | 597 | 141 | 23.6 | 1.39 | 1.07 |
| **Relationship status** | | | | | |
| Not in a relationship | 959 | 159 | 16.6 | 1 | |
| In a relationship | 905 | 212 | 23.4 | 1.54 | 1.22 |
| Ever married/lived with someone | 79 | 21 | 26.6 | 1.82 | 1.08 |
| Unknown | 267 | 113 | 42.3 | 3.69 | 2.74 |
| **Ever had sex** | | | | | |
| No | 864 | 48 | 5.6 | 1 | |
| Yes | 1,008 | 340 | 33.7 | 8.65 | 6.29 |
| Prefer not to say/unknown | 338 | 117 | 34.6 | 9.00 | 6.23 |
| **Called a helpline or searched for information on HIV on the internet** | | | | | |
| No | 1,182 | 171 | 14.5 | 1 | |
| Yes | 930 | 299 | 32.2 | 2.80 | 2.26 |
| Unknown | 98 | 35 | 35.7 | 3.28 | 2.11 |

†age-group specific Odds Ratios (OR); ‡only variables associated with the outcome at p<=0.1 were

### uga campaign, among 15-19 year-olds (A) and 20-24 year-olds (B)

| South 2 | | | | Exposure to any MTV Sh | | | | |
|---|---|---|---|---|---|---|---|---|
| OR† | Fully Adjusted OR‡ | | | Total | Outcome | Row % Outcome | Age Adjusted ( | |
| % CI) | aOR | (95% CI) | | | | | | |
| | | | | 988 | 527 | 53.3 | | |
| | 1 | | | 337 | 156 | 46.3 | 1 | |
| 2.3 | 1.62 | 1.17 | 2.24 | 601 | 360 | 59.9 | 1.73 | 1.32 |
| 0.9 | 1.01 | 0.34 | 3.03 | 50 | 11 | 22 | 0.33 | 0.16 |
| | 1 | | | 536 | 328 | 61.2 | 1 | |
| 0.55 | 0.31 | 0.19 | 0.53 | 137 | 32 | 23.4 | 0.19 | 0.13 |
| 1.23 | 0.61 | 0.42 | 0.89 | 214 | 131 | 61.2 | 1.00 | 0.72 |
| 0.52 | 0.26 | 0.1 | 0.64 | 55 | 15 | 27.3 | 0.24 | 0.13 |
| 0.82 | 0.42 | 0.17 | 1.01 | 46 | 21 | 45.7 | 0.53 | 0.29 |
| | | | | 88 | 45 | 51.1 | 1 | |
| 1.66 | | | | 729 | 401 | 55 | 1.17 | 0.75 |
| 1.98 | | | | 97 | 53 | 54.6 | 1.15 | 0.65 |
| 1.1 | | | | 74 | 28 | 37.8 | 0.58 | 0.31 |
| | 1 | | | 814 | 453 | 55.7 | 1 | |
| 1.36 | 0.88 | 0.52 | 1.48 | 87 | 47 | 54 | 0.94 | 0.6 |
| 0.53 | 0.39 | 0.18 | 0.83 | 87 | 27 | 31 | 0.36 | 0.22 |
| | | | | 650 | 352 | 54.2 | 1 | |
| 1.18 | | | | 113 | 64 | 56.6 | 1.11 | 0.74 |
| 0.84 | | | | 203 | 98 | 48.3 | 0.79 | 0.58 |
| 1.94 | | | | 22 | 13 | 59.1 | 1.22 | 0.52 |
| | 1 | | | 561 | 302 | 53.8 | 1 | |
| 1.56 | 1.28 | 0.9 | 1.83 | 294 | 155 | 52.7 | 0.96 | 0.72 |
| 4.39 | 3.17 | 1.79 | 5.63 | 67 | 47 | 70.1 | 2.02 | 1.16 |
| 1.58 | 2.01 | 0.98 | 4.12 | 66 | 23 | 34.8 | 0.46 | 0.27 |
| | 1 | | | 406 | 191 | 47 | 1 | |
| 1.25 | 0.75 | 0.51 | 1.1 | 280 | 161 | 57.5 | 1.52 | 1.12 |
| 1.92 | 1.09 | 0.71 | 1.67 | 302 | 175 | 57.9 | 1.55 | 1.15 |

| | | | | | | | | |
|---|---|---|---|---|---|---|---|---|
| | 1 | | | 422 | 208 | 49.3 | 1 | |
| 1.76 | 1.22 | 0.86 | 1.73 | 373 | 222 | 59.5 | 1.51 | 1.14 |
| 2.48 | 2.13 | 1.32 | 3.42 | 193 | 97 | 50.3 | 1.04 | 0.74 |
| | 1 | | | 494 | 224 | 45.3 | 1 | |
| 1.77 | 1.23 | 0.84 | 1.79 | 333 | 192 | 57.7 | 1.64 | 1.24 |
| 3.03 | 6.64 | 2.4 | 18.34 | 161 | 111 | 68.9 | 2.68 | 1.83 |
| | 1 | | | 427 | 187 | 43.8 | 1 | |
| 2.03 | 1.37 | 0.92 | 2.04 | 322 | 216 | 67.1 | 2.62 | 1.94 |
| 1.71 | 0.28 | 0.1 | 0.77 | 239 | 124 | 51.9 | 1.38 | 1.01 |
| | 1 | | | 485 | 204 | 42.1 | 1 | |
| 2.27 | 1.61 | 1.18 | 2.21 | 422 | 294 | 69.7 | 3.16 | 2.4 |
| 1.44 | 1.15 | 0.56 | 2.38 | 81 | 29 | 35.8 | 0.77 | 0.47 |

included in the fully adjusted model; §TVET -Technical and Vocational Education and Training; ¶

| | | | | Exposure to any MTV Shuga campaign | | | | |
|---|---|---|---|---|---|---|---|---|
| OR† | Fully Adjusted OR‡ | | | Total | Outcome | Row % Outcome | Age Adjusted ( | |
| % CI) | aOR | (95% CI) | | | | | | |
| | | | | 2,210 | 931 | 42.1 | | |
| | 1 | | | 896 | 299 | 33.4 | 1 | |
| 1.74 | 1.14 | 0.9 | 1.44 | 1,273 | 624 | 49 | 1.92 | 1.61 |
| 1.48 | 0.75 | 0.26 | 2.16 | 41 | 8 | 19.5 | 0.48 | 0.22 |
| | 1 | | | 117 | 58 | 49.6 | 1 | |
| 0.66 | 0.86 | 0.52 | 1.44 | 784 | 164 | 20.9 | 0.27 | 0.18 |
| 2.3 | 1.51 | 0.93 | 2.43 | 880 | 515 | 58.5 | 1.44 | 0.98 |
| 1.13 | 0.68 | 0.4 | 1.15 | 373 | 176 | 47.2 | 0.91 | 0.6 |
| 1.01 | 0.44 | 0.17 | 1.14 | 56 | 18 | 32.1 | 0.48 | 0.25 |
| | 1 | | | 143 | 88 | 61.5 | 1 | |
| 0.82 | 0.81 | 0.54 | 1.23 | 1,834 | 728 | 39.7 | 0.41 | 0.29 |
| 1.39 | 0.83 | 0.46 | 1.5 | 119 | 63 | 52.9 | 0.7 | 0.43 |
| 0.92 | 0.6 | 0.32 | 1.15 | 114 | 52 | 45.6 | 0.52 | 0.32 |

| | | | | | | | | |
|---|---|---|---|---|---|---|---|---|
| | 1 | | | 1,927 | 779 | 40.4 | 1 | |
| 2.51 | 1.71 | 1.04 | 2.8 | 168 | 105 | 62.5 | 2.46 | 1.77 |
| 1.79 | 1.18 | 0.64 | 2.16 | 115 | 47 | 40.9 | 1.02 | 0.69 |
| | 1 | | | 1,655 | 617 | 37.3 | 1 | |
| 2.18 | 0.81 | 0.51 | 1.28 | 208 | 125 | 60.1 | 2.53 | 1.89 |
| 1.55 | 0.78 | 0.54 | 1.13 | 306 | 167 | 54.6 | 2.02 | 1.58 |
| 2.99 | 1.04 | 0.43 | 2.51 | 41 | 22 | 53.7 | 1.95 | 1.05 |
| | 1 | | | 1,224 | 433 | 35.4 | 1 | |
| 2.58 | 1.74 | 1.36 | 2.23 | 754 | 395 | 52.4 | 2.01 | 1.67 |
| 3.21 | 1.97 | 1.27 | 3.07 | 137 | 74 | 54 | 2.15 | 1.5 |
| 2.00 | 1.13 | 0.62 | 2.06 | 95 | 29 | 30.5 | 0.8 | 0.51 |
| | 1 | | | 770 | 379 | 49.2 | 1 | |
| 0.81 | 0.92 | 0.69 | 1.22 | 768 | 278 | 36.2 | 0.59 | 0.48 |
| 1.1 | 0.91 | 0.66 | 1.26 | 672 | 274 | 40.8 | 0.71 | 0.58 |
| | 1 | | | 768 | 295 | 38.4 | 1 | |
| 2.05 | 1.49 | 1.13 | 1.96 | 845 | 416 | 49.2 | 1.55 | 1.28 |
| 1.8 | 2.16 | 1.52 | 3.08 | 597 | 220 | 36.9 | 0.94 | 0.75 |
| | 1 | | | 959 | 304 | 31.7 | 1 | |
| 1.94 | 0.77 | 0.59 | 1.01 | 905 | 417 | 46.1 | 1.84 | 1.52 |
| 3.09 | 0.75 | 0.42 | 1.33 | 79 | 40 | 50.6 | 2.21 | 1.39 |
| 4.97 | 2.96 | 1.57 | 5.58 | 267 | 170 | 63.7 | 3.78 | 2.84 |
| | 1 | | | 864 | 99 | 11.5 | 1 | |
| 11.90 | 7.98 | 5.42 | 11.74 | 1,008 | 652 | 64.7 | 14.15 | 11.07 |
| 13.00 | 3.02 | 1.51 | 6.03 | 338 | 180 | 53.3 | 8.8 | 6.53 |
| | 1 | | | 1,182 | 291 | 24.6 | 1 | |
| 3.47 | 1.22 | 0.95 | 1.57 | 930 | 594 | 63.9 | 5.41 | 4.48 |
| 5.12 | 1.74 | 0.98 | 3.08 | 98 | 46 | 46.9 | 2.71 | 1.78 |

e included in the fully adjusted model; [§]TVET -Technical and Vocational Education and Training; [¶]

| uga campaign | | | |
|---|---|---|---|
| OR† | Fully Adjusted OR‡ | | |
| | aOR | (95% CI) | |
| | 1 | | |
| 2.27 | 1.61 | 1.18 | 2.2 |
| 0.66 | 1.01 | 0.41 | 2.47 |
| | 1 | | |
| 0.30 | 0.18 | 0.11 | 0.29 |
| 1.39 | 0.62 | 0.42 | 0.91 |
| 0.44 | 0.25 | 0.12 | 0.5 |
| 0.98 | 0.83 | 0.38 | 1.81 |
| | 1 | | |
| 1.82 | 1.1 | 0.65 | 1.88 |
| 2.05 | 1.08 | 0.55 | 2.13 |
| 1.09 | 0.56 | 0.27 | 1.15 |
| | 1 | | |
| 1.46 | 1.04 | 0.62 | 1.74 |
| 0.58 | 0.62 | 0.34 | 1.14 |
| 1.65 | | | |
| 1.08 | | | |
| 2.90 | | | |
| | 1 | | |
| 1.27 | 0.96 | 0.68 | 1.36 |
| 3.49 | 2.42 | 1.27 | 4.61 |
| 0.78 | 0.92 | 0.45 | 1.86 |
| | 1 | | |
| 2.07 | 1.29 | 0.88 | 1.88 |
| 2.09 | 1.47 | 0.96 | 2.25 |

| | | | |
|---|---|---|---|
| | 1 | | |
| 2.00 | 1.2 | 0.86 | 1.69 |
| 1.46 | 1.07 | 0.67 | 1.7 |
| | 1 | | |
| 2.17 | 1.31 | 0.91 | 1.89 |
| 3.9 | 6.82 | 3.11 | 14.92 |
| | 1 | | |
| 3.53 | 2.02 | 1.37 | 2.97 |
| 1.9 | 0.38 | 0.18 | 0.81 |
| | 1 | | |
| 4.16 | 2.69 | 1.97 | 3.68 |
| 1.25 | 1.04 | 0.51 | 2.11 |

includes 4 respondents whose language was unknow

| OR† | **Fully Adjusted OR‡** | | |
|---|---|---|---|
| | **aOR** | **(95% CI)** | |
| | 1 | | |
| 2.29 | 1.63 | 1.31 | 2.03 |
| 1.06 | 0.68 | 0.27 | 1.71 |
| | 1 | | |
| 0.4 | 0.48 | 0.3 | 0.76 |
| 2.11 | 1.33 | 0.84 | 2.11 |
| 1.38 | 0.7 | 0.43 | 1.14 |
| 0.94 | 0.44 | 0.21 | 0.96 |
| | 1 | | |
| 0.58 | 0.6 | 0.39 | 0.92 |
| 1.15 | 0.6 | 0.33 | 1.08 |
| 0.86 | 0.54 | 0.3 | 0.99 |

| | | | |
|---|---|---|---|
| | 1 | | |
| 3.4 | 1.01 | 0.62 | 1.64 |
| 1.49 | 0.47 | 0.27 | 0.84 |
| | 1 | | |
| 3.4 | 1.71 | 1.1 | 2.66 |
| 2.59 | 1.68 | 1.17 | 2.39 |
| 3.63 | 2.75 | 1.14 | 6.64 |
| | 1 | | |
| 2.42 | 1.58 | 1.25 | 2.01 |
| 3.06 | 2.14 | 1.36 | 3.36 |
| 1.26 | 0.64 | 0.37 | 1.12 |
| | 1 | | |
| 0.72 | 0.95 | 0.73 | 1.24 |
| 0.88 | 0.91 | 0.67 | 1.25 |
| | 1 | | |
| 1.9 | 1.42 | 1.1 | 1.84 |
| 1.17 | 1.61 | 1.15 | 2.26 |
| | 1 | | |
| 2.22 | 0.79 | 0.61 | 1.02 |
| 3.51 | 0.59 | 0.34 | 1.01 |
| 5.02 | 2.84 | 1.57 | 5.13 |
| | 1 | | |
| 18.1 | 8.31 | 6.1 | 11.31 |
| 11.87 | 2.59 | 1.42 | 4.73 |
| | 1 | | |
| 6.53 | 2.2 | 1.74 | 2.78 |
| 4.11 | 1.75 | 0.99 | 3.11 |

includes 1 respondent whose language was unknown

**Table S2. Associations between exposure to MTV Down South 2 (A) and any MTV Shuga campaig**

| | Exposed | Total | Outcome | Row % Outcome |
|---|---|---|---|---|
| | | N | n | % |
| *(A) MTV Down South 2* | | | | |
| HIV Status | | | | |
| Knowledge of HIV status (N=3,198) | No | 2,393 | 828 | 34.6 |
| | Yes | 805 | 466 | 57.9 |
| HIV Self Testing/Screening outcomes | | | | |
| Ever heard about HIV Self Screening Kit (N=2,727) | No | 2,077 | 589 | 28.4 |
| | Yes | 650 | 387 | 59.5 |
| Ever tested for HIV using HIV Self Screening Kit (N=2,694) | No | 2,049 | 211 | 10.3 |
| | Yes | 645 | 186 | 28.8 |
| Tested for HIV using HIV Self Screening Kit in the last 12 months (N=2,689) | No | 2,044 | 132 | 6.5 |
| | Yes | 645 | 136 | 21.1 |
| Demand for HIV Self Screening - to screen oneself (N=2,213) | No | 1,786 | 1,498 | 83.9 |
| | Yes | 427 | 335 | 78.5 |
| Demand for HIV Self Screening - willing to give kit to partner (N=2,188) | No | 1,774 | 1,502 | 84.7 |
| | Yes | 414 | 330 | 79.7 |
| PrEP Outcomes | | | | |
| Ever Heard About PrEP (N=2,649) | No | 2,023 | 548 | 27.1 |
| | Yes | 626 | 323 | 51.6 |
| Demand for PrEP - willing to take PrEP everyday (N=2,284) | No | 1,747 | 1,394 | 79.8 |
| | Yes | 537 | 457 | 85.1 |
| Sexual behaviour outcomes | | | | |
| Ever had sex (N=2,621) | No | 2,006 | 877 | 43.7 |
| | Yes | 615 | 453 | 73.7 |
| Had sex in the past 12 months (N=2,492) | No | 1,918 | 717 | 37.4 |
| | Yes | 574 | 366 | 63.8 |
| Condom use at last sex with current or last partner (in the last 12 months) (N=1,056) | No | 699 | 410 | 58.7 |
| | Yes | 357 | 234 | 65.5 |
| *(B) Any MTV Shuga campaign* | | | | |
| HIV status | | | | |
| Knowledge of HIV status (N=3,198) | No | 1,740 | 431 | 24.8 |
| | Yes | 1,458 | 863 | 59.2 |
| HIV Self Testing/Screening outcomes | | | | |
| Ever heard about HIV Self Screening Kit (N=2,727) | No | 1,509 | 284 | 18.8 |
| | Yes | 1,218 | 692 | 56.8 |
| Ever tested for HIV using HIV Self Screening Kit (N=2,694) | No | 1,483 | 115 | 7.8 |
| | Yes | 1,211 | 282 | 23.3 |
| Tested for HIV using HIV Self Screening Kit in the last 12 months (N=2,689) | No | 1,478 | 70 | 4.7 |
| | Yes | 1,211 | 198 | 16.4 |
| Demand for HIV Self Screening - to screen oneself (N=2,213) | No | 1,342 | 1,143 | 85.2 |
| | Yes | 871 | 690 | 79.2 |
| Demand for HIV Self Screening - willing to give kit to partner (N=2,188) | No | 1,338 | 1,145 | 85.6 |
| | Yes | 850 | 687 | 80.8 |
| PrEP Outcomes | | | | |

| | | | | |
|---|---|---|---|---|
| **Ever Heard About PrEP (N=2,649)** | No | 1,469 | 251 | 17.1 |
| | Yes | 1,180 | 620 | 52.5 |
| **Demand for PrEP - willing to take PrEP everyday (N=2,284)** | No | 1,292 | 1,011 | 78.3 |
| | Yes | 992 | 840 | 84.7 |
| **Sexual behaviour outcomes** | | | | |
| **Ever had sex (N=2,621)** | No | 1,467 | 462 | 31.5 |
| | Yes | 1,154 | 868 | 75.2 |
| **Had sex in the past 12 months (N=2,492)** | No | 1,419 | 377 | 26.6 |
| | Yes | 1,073 | 706 | 65.8 |
| **Condom use at last sex with current or last partner (in the last 12 months) (N=1,056)** | No | 365 | 206 | 56.4 |
| | Yes | 691 | 438 | 63.4 |

[†]all models (except models on sexual behaviours) include the following variables: main exposure, age, gender, schooling, [illegible] status, ever had sex, and exposure to other non-MTV resources; models for sexual behaviours include all the above variab[illegible]

### n (B) with HIV self-testing and PrEP outcomes: Age specific estimates

| Fully adjusted† OR (overall) | | | | Fully adjusted† age specific Odds Ratios (OR): 15-19 year olds | | | | 20-24 year olds | | | |
|---|---|---|---|---|---|---|---|---|---|---|---|
| aOR | (95% CI) | | p value | aOR | (95% CI) | | p value | aOR | (95% CI) | | p value |
| 1 | | | | 1 | | | | 1 | | | |
| 2.06 | 1.64 | 2.58 | <0.001 | 1.60 | 1.12 | 2.30 | 0.010 | 2.61 | 1.97 | 3.47 | <0.001 |
| 1 | | | | 1 | | | | 1 | | | |
| 1.99 | 1.61 | 2.47 | <0.001 | 1.87 | 1.31 | 2.68 | 0.001 | 2.96 | 2.29 | 3.84 | <0.001 |
| 1 | | | | | | | | | | | |
| 2.49 | 1.95 | 3.19 | <0.001 | No evidence of age/DS2 interaction (p=0.160) | | | | | | | |
| 1 | | | | | | | | | | | |
| 2.61 | 1.97 | 3.47 | <0.001 | No evidence of age/DS2 interaction (p=0.219) | | | | | | | |
| 1 | | | | 1 | | | | 1 | | | |
| 0.80 | 0.59 | 1.09 | 0.156 | 1.66 | 1.05 | 2.64 | 0.031 | 0.50 | 0.34 | 0.72 | <0.001 |
| 1 | | | | 1 | | | 1 | | | | |
| 0.86 | 0.63 | 1.17 | 0.326 | 1.84 | 1.13 | 3.00 | 0.014 | 0.52 | 0.36 | 0.77 | 0.001 |
| 1 | | | | | | | | | | | |
| 1.90 | 1.53 | 2.35 | <0.001 | No evidence of age/DS2 interaction (p=0.764) | | | | | | | |
| 1 | | | | | | | | | | | |
| 1.31 | 0.97 | 1.76 | 0.079 | Weak evidence of age/DS2 interaction (p=0.080) | | | | | | | |
| 1 | | | | 1 | | | | 1 | | | |
| 2.86 | 2.19 | 3.72 | <0.001 | 1.49 | 1.02 | 2.18 | 0.038 | 8.05 | 5.57 | 11.65 | <0.001 |
| 1 | | | | 1 | | | | 1 | | | |
| 2.13 | 1.65 | 2.74 | <0.001 | 1.39 | 0.94 | 2.06 | 0.098 | 4.24 | 3.08 | 5.82 | <0.001 |
| 1 | | | | | | | | | | | |
| 1.42 | 1.07 | 1.89 | 0.014 | No evidence of age/DS2 interaction (p=0.503) | | | | | | | |
| 1 | | | | 1 | | | | 1 | | | |
| 2.52 | 2.04 | 3.12 | <0.001 | 1.85 | 1.32 | 2.60 | <0.001 | 3.86 | 3.01 | 4.95 | <0.001 |
| 1 | | | | | | | | | | | |
| 3.00 | 2.45 | 3.68 | <0.001 | No evidence of age/MTV interaction (p=0.123) | | | | | | | |
| 1 | | | | | | | | | | | |
| 2.15 | 1.64 | 2.81 | <0.001 | No evidence of age/MTV interaction (p=0.429) | | | | | | | |
| 1 | | | | | | | | | | | |
| 2.23 | 1.61 | 3.07 | <0.001 | No evidence of age/MTV interaction (p=0.128) | | | | | | | |
| 1 | | | | 1 | | | | 1 | | | |
| 0.73 | 0.55 | 0.97 | 0.031 | 1.55 | 1.03 | 2.34 | 0.034 | 0.49 | 0.34 | 0.69 | <0.001 |
| 1 | | | | 1 | | | | 1 | | | |
| 0.87 | 0.65 | 1.17 | 0.364 | 1.90 | 1.24 | 2.90 | 0.003 | 0.58 | 0.40 | 0.82 | 0.003 |

| | | | | | | | | | | | |
|---|---|---|---|---|---|---|---|---|---|---|---|
| 1 | | | | | | | | | | | |
| 2.8 | 2.26 | 3.47 | <0.001 | No evidence of age/MTV interaction (p=0.834) | | | | | | | |
| 1 | | | | 1 | | | | 1 | | | |
| 1.26 | 0.96 | 1.65 | 0.095 | 2.39 | 1.58 | 3.60 | <0.001 | 0.99 | 0.72 | 1.37 | 0.960 |
| 1 | | | | 1 | | | | 1 | | | |
| 4.29 | 3.36 | 5.48 | <0.001 | 2.46 | 1.71 | 3.55 | <0.001 | 11.38 | 8.49 | 15.27 | <0.001 |
| 1 | | | | 1 | | | | 1 | | | |
| 3.03 | 2.39 | 3.84 | <0.001 | 2.14 | 1.46 | 3.14 | <0.001 | 6.44 | 4.93 | 8.41 | <0.001 |
| 1 | | | | | | | | | | | |
| 1.61 | 1.21 | 2.13 | 0.001 | No evidence of age/MTV interaction (p=0.142) | | | | | | | |

ome language, rural/urban setting, province, food insecurity, household media index, individual media index, relationship
les except ever had sex.

**Table S3. Results from multiple imputation with primary exposure (Down South 2): m=10 imputati**
All variables included in the complete case analysis were included in the imputation models

| | |
|---|---|
| **HIV status** | **Knowledge of HIV status (N=3,431)** |
| **HIV Self-Screening/Testing outcomes** | **Ever heard about HIV Self Screening Kit (N=3,431)** |
| | **Ever tested for HIV using HIV Self Screening Kit (N=3,431)** |
| | **Tested for HIV using HIV Self Screening Kit in the last 12 m** |
| | **Demand for HIVSS - to screen oneself (N=3,034)** |
| | **Demand for HIVSS - willing to give kit to partner (N=3,034)** |
| **PrEP Outcomes** | **Ever Heard About PrEP (N=3,431)** |
| | **Demand for PrEP - willing to take PrEP everyday (N=3,431)** |
| **Sexual behaviour outcome** | **Ever had sex (N=3,431)** |
| | **Had sex in the past 12 months (N=3,431)** |
| | **Condom use at last sex with current or last partner (in the l** |

ons

| Age Adjusted OR | | | p value | Fully adjusted OR | | | p value (LR test) | Fully adjusted OR for 15-19 yr olds | | | p value (LR test) |
|---|---|---|---|---|---|---|---|---|---|---|---|
| aOR | (95% CI) | | | aOR | (95% CI) | | aOR | aOR | (95% CI) | | |
| 1 | | | | 1 | | | | | | | |
| 2.93 | 2.49 | 3.45 | <0.001 | 1.70 | 1.39 | 2.09 | <0.001 | *No evidence of age/DS2 interactic | | | |
| 1 | | | | 1 | | | | 1 | | | |
| 3.64 | 3.03 | 4.36 | <0.001 | 2.46 | 2.01 | 3.02 | <0.001 | 1.73 | 1.27 | 2.34 | <0.001 |
| 1 | | | | 1 | | | | | | | |
| 3.53 | 2.83 | 4.42 | <0.001 | 2.51 | 1.96 | 3.21 | <0.001 | *No evidence of age/DS2 interactic | | | |
| 1 | | | | 1 | | | | | | | |
| 4.00 | 3.09 | 5.20 | <0.001 | 2.56 | 1.94 | 3.39 | <0.001 | *No evidence of age/DS2 interactic | | | |
| 1 | | | | 1 | | | | 1 | | | |
| 0.79 | 0.61 | 1.03 | 0.086 | 0.78 | 0.56 | 1.08 | 0.127 | 1.24 | 0.80 | 1.94 | 0.327 |
| 1 | | | | 1 | | | | 1 | | | |
| 0.85 | 0.60 | 1.20 | 0.343 | 0.87 | 0.59 | 1.28 | 0.468 | 1.43 | 0.94 | 2.19 | 0.097 |
| 1 | | | | 1 | | | | | | | |
| 2.81 | 2.35 | 3.36 | <0.001 | 1.86 | 1.49 | 2.32 | <0.001 | *No evidence of age/DS2 interactic | | | |
| 1 | | | | 1 | | | | | | | |
| 1.62 | 1.17 | 2.24 | 0.005 | 1.37 | 0.94 | 1.99 | 0.098 | *No evidence of age/DS2 interactic | | | |
| 1 | | | | 1 | | | | 1 | | | |
| 3.46 | 2.88 | 4.17 | <0.001 | 2.87 | 2.22 | 3.70 | <0.001 | 1.10 | 0.77 | 1.57 | 0.599 |
| 1 | | | | 1 | | | | 1 | | | |
| 2.72 | 2.23 | 3.31 | <0.001 | 2.10 | 1.64 | 2.68 | <0.001 | 0.98 | 0.66 | 1.45 | 0.914 |
| 1 | | | | 1 | | | | | | | |
| 1.69 | 1.29 | 2.21 | <0.001 | 1.80 | 1.37 | 2.36 | <0.001 | *No evidence of age/DS2 interactic | | | |

| Fully adjusted OR for 20-24 yr olds | | | p value (LR test) |
|---|---|---|---|
| aOR | (95% CI) | | |
| on (p=0.600) | | | |
| 1 | | | |
| 2.91 | 2.28 | 3.72 | <0.001 |
| on (p=0.202) | | | |
| on (p=0.114) | | | |
| 1 | | | |
| 0.52 | 0.35 | 0.76 | <0.001 |
| 1 | | | |
| 0.58 | 0.41 | 0.83 | 0.003 |
| on (p=0.776) | | | |
| on (p=0.144) | | | |
| 1 | | | |
| 6.48 | 4.37 | 9.63 | <0.001 |
| 1 | | | |
| 3.26 | 2.37 | 4.49 | <0.001 |
| on (p=0.327 ) | | | |

**Table S4. Experiences with HIV self-testing and PrEP, among those ever and never exposed to MT**

| | 15-19 year olds | | | |
|---|---|---|---|---|
| | **Unexposed to DS2** | | **Exposed to DS2** | |
| | N | % | N | % |
| **HIV testing experiences with community health worker (CHW)** | | | | |
| How many times ever tested (with a CHW) | | | | |
| 1 | 31 | 11.9 | 22 | 16.1 |
| 2 | 31 | 11.9 | 18 | 13.1 |
| 3-5 | 83 | 31.9 | 43 | 31.4 |
| 6-10 | 36 | 13.8 | 24 | 17.5 |
| 11+ | 20 | 7.7 | 13 | 9.5 |
| Missing | 59 | 22.7 | 17 | 12.4 |
| **Total** | **260** | **100** | **137** | **100** |
| How many times tested in past 12 months (with a CHW) | | | | |
| 1 | 39 | 15 | 22 | 16.1 |
| 2 | 44 | 16.9 | 27 | 19.7 |
| 3-5 | 62 | 23.8 | 38 | 27.7 |
| 6-10 | 12 | 4.6 | 8 | 5.8 |
| 11+ | 0 | 0 | 1 | 0.7 |
| Missing | 103 | 39.6 | 41 | 29.9 |
| **Total** | **260** | **100** | **137** | **100** |
| When was the last time you tested for HIV? | | | | |
| In the last year | 26 | 10 | 9 | 6.6 |
| 1 to 2 years ago | 25 | 9.6 | 19 | 13.9 |
| More than 3 years ago | 18 | 6.9 | 4 | 2.9 |
| Do not know | 2 | 0.8 | 0 | 0 |
| Prefer not to say | 4 | 1.5 | 0 | 0 |
| Missing | 185 | 71.2 | 105 | 76.6 |
| **Total** | **260** | **100** | **137** | **100** |
| Was your last HIV test offered during an ante-natal care visit? | | | | |
| No | 29 | 11.2 | 19 | 13.9 |
| Yes | 24 | 9.2 | 20 | 14.6 |
| I have never been pregnant (non applicable) | 88 | 33.8 | 50 | 36.5 |
| Do not know | 10 | 3.8 | 6 | 4.4 |
| Prefer not to say | 4 | 1.5 | 2 | 1.5 |
| Missing | 105 | 40.4 | 40 | 29.2 |
| **Total** | **260** | **100** | **137** | **100** |
| The last time you tested for HIV with a health worker, where did you do your test | | | | |
| Government hospital | 63 | 24.2 | 30 | 21.9 |
| Government clinic/ community health centre | 115 | 44.2 | 57 | 41.6 |
| University/TVET/school clinic | 23 | 8.8 | 19 | 13.9 |
| Mobile HIV testing services | 24 | 9.2 | 15 | 10.9 |
| New Start testing site | 2 | 0.8 | 1 | 0.7 |
| Pharmacy/ chemist | 3 | 1.2 | 7 | 5.1 |
| Private hospital/clinic/doctor | 21 | 8.1 | 6 | 4.4 |
| Other: Specify | 6 | 2.3 | 0 | 0 |
| Do not know | 2 | 0.8 | 1 | 0.7 |
| Prefer not to say | 1 | 0.4 | 1 | 0.7 |
| Missing | | | | |
| **Total** | **260** | **100** | **137** | **100** |
| Reasons for the last test | | | | |
| I wanted to know my HIV status | 201 | 77.3 | 119 | 86.9 |
| I heard about HIV testing in MTV Shuga Down Sou | 11 | 4.2 | 26 | 19 |
| I heard about HIV test in another TV/radio broadca | 5 | 1.9 | 16 | 11.7 |

| | | | | |
|---|---|---|---|---|
| I had sex without a condom | 38 | 14.6 | 26 | 19 |
| I was sick and worried | 8 | 3.1 | 14 | 10.2 |
| I was tested while pregnant | 11 | 4.2 | 8 | 5.8 |
| I was referred for HIV testing by a Health Care Wor | 15 | 5.8 | 15 | 10.9 |
| I had to confirm my HIV status to get PrEP | 7 | 2.7 | 7 | 5.1 |
| My partner/husband wanted to know my HIV status | 6 | 2.3 | 3 | 2.2 |
| My partner tested positive | 1 | 0.4 | 1 | 0.7 |
| My client wanted to know my HIV status | 0 | | | |
| The migration office required my HIV status | 1 | 0.4 | 0 | 0 |
| My employer required my HIV status | 1 | 0.4 | 0 | 0 |
| I wanted to get circumcised | 27 | 10.4 | 11 | 8 |
| **Total** | **260** | **100.0** | **137** | **100** |

**HIV Self-Screening (HIV SS) experiences**

| How many times ever used a Self-Screening kit | | | | |
|---|---|---|---|---|
| 1 | 15 | 31.9 | 7 | 12.5 |
| 2 | 8 | 17 | 10 | 17.9 |
| 3-5 | 5 | 10.6 | 14 | 25 |
| 6-10 | 7 | 14.9 | 10 | 17.9 |
| 11+ | 3 | 6.4 | 9 | 16.1 |
| Missing | 9 | 19.1 | 6 | 10.7 |
| **Total** | **47** | **100** | **56** | **100** |

| How many times tested in past 12 months with a HIV Self Screening (HIV SS) kit | | | | |
|---|---|---|---|---|
| 1 | 7 | 14.9 | 5 | 8.9 |
| 2 | 7 | 14.9 | 13 | 23.2 |
| 3-5 | 5 | 10.6 | 7 | 12.5 |
| 6-10 | 1 | 2.1 | 8 | 14.3 |
| 11+ | 1 | 2.1 | 1 | 1.8 |
| Missing | 26 | 55.3 | 22 | 39.3 |
| **Total** | **47** | **100** | **56** | **100** |

| When was the last time you used a HIV Self-Screening kit? | | | | |
|---|---|---|---|---|
| In the last year | 10 | 21.3 | 7 | 12.5 |
| 1 to 2 year ago | 4 | 8.5 | 7 | 12.5 |
| More than 3 years ago | 2 | 4.3 | 4 | 7.1 |
| Do not know | 1 | 2.1 | 2 | 3.6 |
| Prefer not to say | 2 | 4.3 | 0 | 0 |
| Missing | 28 | 59.6 | 36 | 64.3 |
| **Total** | **47** | **100** | **56** | **100** |

| Was your last HIV Self-Screening test offered during an ante-natal care visit? | | | | |
|---|---|---|---|---|
| No | 6 | 12.8 | 13 | 23.2 |
| Yes | 5 | 10.6 | 11 | 19.6 |
| I have never been pregnant (non applicable) | 14 | 29.8 | 14 | 25 |
| Do not know | 1 | 2.1 | 0 | 0 |
| Prefer not to say | 1 | 2.1 | 0 | 0 |
| Missing | 20 | 42.6 | 18 | 32.1 |
| **Total** | **47** | **100** | **56** | **100** |

| The last time you used a HIV Self-Screening test, where did you get the kit from | | | | |
|---|---|---|---|---|
| Government hospital | 7 | 14.9 | 10 | 17.9 |
| Government clinic/ community health centre | 16 | 34 | 16 | 28.6 |
| University / TVET / school | 6 | 12.8 | 7 | 12.5 |
| Mobile HIV testing services | 4 | 8.5 | 4 | 7.1 |
| New Start testing site | 1 | 2.1 | 1 | 1.8 |
| Pharmacy/ chemist | 8 | 17 | 12 | 21.4 |
| Private hospital/clinic | 3 | 6.4 | 5 | 8.9 |
| Other: Specify | 0 | 0 | 1 | 1.8 |
| Do not know | 2 | 4.3 | 0 | 0 |
| Prefer not to say | | | | |

| | | | | |
|---|---|---|---|---|
| Missing | | | | |
| **Total** | **47** | **100** | **56** | **100** |
| The last time you used a HIV Self-Screening test, who did you obtain the kit from | | | | |
| Health care worker | 25 | 53.2 | 20 | 35.7 |
| Community health care worker | 7 | 14.9 | 12 | 21.4 |
| Community-based distribution agent | 2 | 4.3 | 2 | 3.6 |
| Voluntary Medical Male Circumcision agent | 1 | 2.1 | 2 | 3.6 |
| Pharmacist/ chemist | 8 | 17 | 15 | 26.8 |
| Private doctor | 1 | 2.1 | 1 | 1.8 |
| Friend | 0 | 0 | 2 | 3.6 |
| Partner/Spouse | | | | |
| Family member | 0 | 0 | 1 | 1.8 |
| Other: Specify | | | | |
| Do not know | 2 | 4.3 | 1 | 1.8 |
| Prefer not to say | 1 | 2.1 | 0 | 0 |
| Missing | | | | |
| **Total** | **47** | **45.6** | **56** | **54.4** |
| Did you have to pay any fees to get the Self-Screening kit? | | | | |
| No | 32 | 68.1 | 36 | 64.3 |
| Yes | 11 | 23.4 | 20 | 35.7 |
| Do not know | 2 | 4.3 | 0 | 0 |
| Prefer not to say | 2 | 4.3 | 0 | 0 |
| **Total** | **47** | **100** | **56** | **100** |
| How much did you pay to get the Self-Screening kit? | | | | |
| Between R1,00 and R50 | 4 | 8.5 | 7 | 12.5 |
| Between R51,00 and R100 | 3 | 6.4 | 6 | 10.7 |
| Between R101,00 and R150 | 0 | 0 | 4 | 7.1 |
| Between R151,00 and R200 | 2 | 4.3 | 0 | 0 |
| More than R201,00 | 1 | 2.1 | 0 | 0 |
| Do not know | 1 | 2.1 | 3 | 5.4 |
| Prefer not to say | | | | |
| Missing | 36 | 76.6 | 36 | 64.3 |
| **Total** | **47** | **100** | **56** | **100** |
| The last time you used a HIV Self-Screening test, where did you do use your Self-Screening kit | | | | |
| Ante-natal care visit | 1 | 2.1 | 1 | 1.8 |
| Government hospital (not ante-natal care) | 11 | 23.4 | 10 | 17.9 |
| Government clinic/ community health centre (not ar | 9 | 19.1 | 10 | 17.9 |
| University / TVET / school | 7 | 14.9 | 6 | 10.7 |
| Mobile HIV testing services | 4 | 8.5 | 3 | 5.4 |
| New Start testing site | 0 | 0 | 3 | 5.4 |
| Pharmacy/ chemist | 1 | 2.1 | 3 | 5.4 |
| Private hospital/clinic/doctor | 2 | 4.3 | 0 | 0 |
| Home | 11 | 23.4 | 17 | 30.4 |
| Friends home | | | | |
| Partner/Spouses home | | | | |
| Do not know | 1 | 2.1 | 1 | 1.8 |
| Prefer not to say | 0 | 0 | 2 | 3.6 |
| Missing | | | | |
| **Total** | **47** | **100** | **56** | **100** |
| Reason for last SS test | | | | |
| I wanted to know my HIV status | 40 | 85.1 | 45 | 80.4 |
| I heard about HIV self-screening in MTV Shuga Dc | 3 | 6.4 | 18 | 32.1 |
| I heard about HIV self-screening in another TV/rad | 4 | 8.5 | 11 | 19.6 |
| I had sex without a condom | 12 | 25.5 | 11 | 19.6 |
| I was sick and worried | 4 | 8.5 | 10 | 17.9 |
| I was tested while pregnant | 2 | 4.3 | 3 | 5.4 |

| | | | | |
|---|---|---|---|---|
| I was referred for HIV self-screening by a Health Ca | 1 | 2.1 | 10 | 17.9 |
| I had to confirm my HIV status to get PrEP | 1 | 2.1 | 4 | 7.1 |
| My partner/husband wanted to know my HIV status | 1 | 2.1 | 2 | 3.6 |
| My partner tested positive | 0 | 0 | 1 | 1.8 |
| My client wanted to know my HIV status | 0 | | | |
| The migration office required my HIV status | 1 | 2.1 | 0 | 0 |
| My employer required my HIV status | 2 | 4.3 | 0 | 0 |
| I wanted to get circumcised | 1 | 2.1 | 5 | 8.9 |
| **Total** | **47** | **100** | **56** | **100** |
| Did you go to a health facility to confirm the result of your HIV Self-Screening through a lab test? | | | | |
| No | 20 | 42.6 | 19 | 33.9 |
| Yes | 26 | 55.3 | 37 | 66.1 |
| Do not know | | | | |
| Prefer not to say | 1 | 2.1 | 0 | 0 |
| Missing | | | | |
| **Total** | **47** | **100** | **56** | **100** |
| **Knowledge of HIV Self-Screening (HIV SS)** | | | | |
| Where (or from whom) have you heard about HIV SS | | | | |
| By a health care worker | 91 | 38.9 | 75 | 67.6 |
| By a community health care worker | 35 | 15 | 24 | 21.6 |
| By a community-based distribution agent | 15 | 6.4 | 13 | 11.7 |
| By a Voluntary Medical Male Circumcision agent | 18 | 7.7 | 8 | 7.2 |
| By a family member | 30 | 12.8 | 32 | 28.8 |
| By a friend | 55 | 23.5 | 31 | 27.9 |
| By a peer educator at workplace | 9 | 3.8 | 12 | 10.8 |
| By a peer educator at school/university/TVET | 33 | 14.1 | 30 | 27 |
| By a professional educator or teacher at school/un | 12 | 5.1 | 17 | 15.3 |
| By watching TV | 27 | 11.5 | 33 | 29.7 |
| By listening to radio | 10 | 4.3 | 20 | 18 |
| By reading a graphic novel or magazine | 3 | 1.3 | 11 | 9.9 |
| On the internet or social media | 33 | 14.1 | 21 | 18.9 |
| During a community event | 4 | 1.7 | 8 | 7.2 |
| During a national event | 4 | 1.7 | 1 | 0.9 |
| **Total** | **234** | **100** | **111** | **100** |
| What program told you about HIV SS | | | | |
| MTV Shuga Down South | 10 | 19.6 | 24 | 60 |
| B-wise | 6 | 11.8 | 6 | 15 |
| PrEPWatch | 5 | 9.8 | 6 | 15 |
| MyPrEP | 2 | 3.9 | 5 | 12.5 |
| SheConquers | 1 | 2 | 5 | 12.5 |
| DREAMS | 2 | 3.9 | 4 | 10 |
| Lovelife | 14 | 27.5 | 15 | 37.5 |
| **Total** | **51** | **100** | **40** | **100** |
| Do you know where to go to get an HIVSS kit? | | | | |
| No | 457 | 73.5 | 146 | 59.8 |
| Yes | 109 | 17.5 | 93 | 38.1 |
| Prefer not to say | 55 | 8.8 | 5 | 2 |
| Missing | 1 | 0.2 | 0 | 0 |
| **Total** | **622** | **100** | **244** | **100** |
| **Among those who had never tested (N=1,142)** | | | | |
| Reason why never tested | | | | |
| I have never thought about testing for HIV | 140 | 51.5 | 44 | 50 |
| I'm not at risk of being HIV positive or contracting H | 95 | 34.9 | 17 | 19.3 |
| I'm too young to test | 57 | 21 | 15 | 17 |
| I'm not yet sexually active | 101 | 37.1 | 51 | 58 |

| | | | | |
|---|---|---|---|---|
| I don't want to know my HIV status | 6 | 2.2 | 1 | 1.1 |
| I don't feel sick enough to test for HIV | 10 | 3.7 | 2 | 2.3 |
| I'm afraid of testing positive | 11 | 4 | 4 | 4.5 |
| I'm afraid of what others will think if I take an HIV te | 3 | 1.1 | 4 | 4.5 |
| My partner won't let me test | 3 | 1.1 | 0 | 0 |
| My family members won't let me test | 2 | 0.7 | 1 | 1.1 |
| I'm afraid of the consequences of testing on my rel | 4 | 1.5 | 1 | 1.1 |
| I'm afraid that my test result will not be kept private | 7 | 2.6 | 1 | 1.1 |
| A HIV test is too expensive | 3 | 1.1 | 0 | 0 |
| HIV treatment is too expensive | 1 | 0.4 | 0 | 0 |
| I don't know where to go or how to get tested | 3 | 1.1 | 2 | 2.3 |
| The place where I can go for testing is too far awa | 3 | 1.1 | 3 | 3.4 |
| I don't have the time to go for testing | 8 | 2.9 | 4 | 4.5 |
| **Total** | **272** | **100** | **88** | **100** |

**V Shuga DS2, by age group**

| 20-24 year olds | | | |
|---|---|---|---|
| **Unexposed to DS2** | | **Exposed to DS2** | |
| N | % | N | % |
| 49 | 6.4 | 10 | 2.7 |
| 95 | 12.5 | 33 | 9.1 |
| 199 | 26.1 | 99 | 27.2 |
| 166 | 21.8 | 103 | 28.3 |
| 111 | 14.6 | 60 | 16.5 |
| 142 | 18.6 | 59 | 16.2 |
| **762** | **100** | **364** | **100** |
| 112 | 14.7 | 25 | 6.9 |
| 132 | 17.3 | 87 | 23.9 |
| 208 | 27.3 | 132 | 36.3 |
| 37 | 4.9 | 25 | 6.9 |
| 4 | 0.5 | 9 | 2.5 |
| 269 | 35.3 | 86 | 23.6 |
| **762** | **100** | **364** | **100** |
| 46 | 6 | 21 | 5.8 |
| 71 | 9.3 | 20 | 5.5 |
| 70 | 9.2 | 7 | 1.9 |
| 6 | 0.8 | 0 | 0 |
| 4 | 0.5 | 0 | 0 |
| 565 | 74.1 | 316 | 86.8 |
| **762** | **100** | **364** | **100** |
| 122 | 16 | 67 | 18.4 |
| 116 | 15.2 | 80 | 22 |
| 159 | 20.9 | 79 | 21.7 |
| 20 | 2.6 | 2 | 0.5 |
| 16 | 2.1 | 4 | 1.1 |
| 329 | 43.2 | 132 | 36.3 |
| **762** | **100** | **364** | **100** |
| 198 | 26 | 88 | 24.2 |
| 296 | 38.8 | 133 | 36.5 |
| 109 | 14.3 | 76 | 20.9 |
| 107 | 14 | 29 | 8 |
| 1 | 0.1 | 1 | 0.3 |
| 15 | 2 | 15 | 4.1 |
| 30 | 3.9 | 20 | 5.5 |
| 4 | 0.5 | 1 | 0.3 |
| 0 | 0 | 1 | 0.3 |
| 2 | 0.3 | 0 | 0 |
| **762** | **100** | **364** | **100** |
| 596 | 78.2 | 300 | 82.4 |
| 21 | 2.8 | 64 | 17.6 |
| 21 | 2.8 | 29 | 8 |

| | | | |
|---|---|---|---|
| 146 | 19.2 | 90 | 24.7 |
| 42 | 5.5 | 22 | 6 |
| 57 | 7.5 | 31 | 8.5 |
| 25 | 3.3 | 21 | 5.8 |
| 13 | 1.7 | 14 | 3.8 |
| 15 | 2 | 31 | 8.5 |
| 4 | 0.5 | 3 | 0.8 |
| 4 | 0.5 | 3 | 0.8 |
| 1 | 0.1 | 1 | 0.3 |
| 4 | 0.5 | 1 | 0.3 |
| 80 | 10.5 | 17 | 4.7 |
| **762** | **100** | **364** | **100** |

| | | | |
|---|---|---|---|
| 42 | 25.6 | 28 | 21.5 |
| 30 | 18.3 | 17 | 13.1 |
| 34 | 20.7 | 34 | 26.2 |
| 14 | 8.5 | 16 | 12.3 |
| 10 | 6.1 | 10 | 7.7 |
| 34 | 20.7 | 25 | 19.2 |
| **164** | **100** | **130** | **100** |

| | | | |
|---|---|---|---|
| 35 | 21.3 | 22 | 16.9 |
| 21 | 12.8 | 23 | 17.7 |
| 31 | 18.9 | 32 | 24.6 |
| 4 | 2.4 | 4 | 3.1 |
| 5 | 3 | 5 | 3.8 |
| 68 | 41.5 | 44 | 33.8 |
| **164** | **100** | **130** | **100** |

| | | | |
|---|---|---|---|
| 19 | 11.6 | 9 | 6.9 |
| 20 | 12.2 | 12 | 9.2 |
| 3 | 1.8 | 2 | 1.5 |
| 7 | 4.3 | 3 | 2.3 |
| 5 | 3 | 2 | 1.5 |
| 110 | 67.1 | 102 | 78.5 |
| **164** | **100** | **130** | **100** |

| | | | |
|---|---|---|---|
| 30 | 18.3 | 13 | 10 |
| 24 | 14.6 | 39 | 30 |
| 36 | 22 | 29 | 22.3 |
| 5 | 3 | 1 | 0.8 |
| 5 | 3 | 1 | 0.8 |
| 64 | 39 | 47 | 36.2 |
| **164** | **100** | **130** | **100** |

| | | | |
|---|---|---|---|
| 45 | 27.4 | 32 | 24.6 |
| 48 | 29.3 | 41 | 31.5 |
| 15 | 9.1 | 19 | 14.6 |
| 19 | 11.6 | 9 | 6.9 |
| 1 | 0.6 | 2 | 1.5 |
| 26 | 15.9 | 17 | 13.1 |
| 4 | 2.4 | 7 | 5.4 |
| 1 | 0.6 | 0 | 0 |
| 2 | 1.2 | 2 | 1.5 |
| 2 | 1.2 | 1 | 0.8 |

| | | | |
|---|---|---|---|
| 1 | 0.6 | 0 | 0 |
| **164** | **100** | **130** | **100** |

| | | | |
|---|---|---|---|
| 79 | 48.2 | 73 | 56.2 |
| 34 | 20.7 | 27 | 20.8 |
| 3 | 1.8 | 1 | 0.8 |
| 2 | 1.2 | 0 | 0 |
| 25 | 15.2 | 18 | 13.8 |
| 3 | 1.8 | 1 | 0.8 |
| 6 | 3.7 | 2 | 1.5 |
| 3 | 1.8 | 3 | 2.3 |
| 2 | 1.2 | 1 | 0.8 |
| 1 | 0.6 | 1 | 0.8 |
| 2 | 1.2 | 2 | 1.5 |
| 4 | 2.4 | 0 | 0 |
| 0 | 0 | 1 | 0.8 |
| **164** | **100** | **130** | **100** |

| | | | |
|---|---|---|---|
| 116 | 70.7 | 79 | 60.8 |
| 34 | 20.7 | 47 | 36.2 |
| 6 | 3.7 | 2 | 1.5 |
| 8 | 4.9 | 2 | 1.5 |
| **164** | **100** | **130** | **100** |

| | | | |
|---|---|---|---|
| 18 | 11 | 24 | 18.5 |
| 4 | 2.4 | 10 | 7.7 |
| 5 | 3 | 4 | 3.1 |
| 1 | 0.6 | 1 | 0.8 |
| 1 | 0.6 | 0 | 0 |
| 3 | 1.8 | 4 | 3.1 |
| 2 | 1.2 | 4 | 3.1 |
| 130 | 79.3 | 83 | 63.8 |
| **164** | **100** | **130** | **100** |

| | | | |
|---|---|---|---|
| 5 | 3 | 7 | 5.4 |
| 42 | 25.6 | 30 | 23.1 |
| 28 | 17.1 | 22 | 16.9 |
| 14 | 8.5 | 21 | 16.2 |
| 11 | 6.7 | 5 | 3.8 |
| 1 | 0.6 | 2 | 1.5 |
| 2 | 1.2 | 3 | 2.3 |
| 2 | 1.2 | 4 | 3.1 |
| 51 | 31.1 | 30 | 23.1 |
| 1 | 0.6 | 2 | 1.5 |
| 0 | 0 | 1 | 0.8 |
| 1 | 0.6 | 1 | 0.8 |
| 3 | 1.8 | 1 | 0.8 |
| 3 | 1.8 | 1 | 0.8 |
| **164** | **100** | **130** | **100** |

| | | | |
|---|---|---|---|
| 138 | 84.1 | 113 | 86.9 |
| 5 | 3 | 24 | 18.5 |
| 12 | 7.3 | 15 | 11.5 |
| 28 | 17.1 | 29 | 22.3 |
| 5 | 3 | 7 | 5.4 |
| 11 | 6.7 | 8 | 6.2 |

| | | | |
|---|---|---|---|
| 2 | 1.2 | 5 | 3.8 |
| 5 | 3 | 2 | 1.5 |
| 7 | 4.3 | 9 | 6.9 |
| 0 | 0 | 5 | 3.8 |
| 0 | 0 | 2 | 1.5 |
| 0 | 0 | 3 | 2.3 |
| 0 | 0 | 2 | 1.5 |
| 6 | 3.7 | 3 | 2.3 |
| **164** | **100** | **130** | **100** |

| | | | |
|---|---|---|---|
| 60 | 36.6 | 43 | 33.1 |
| 95 | 57.9 | 86 | 66.2 |
| 2 | 1.2 | 0 | 0 |
| 6 | 3.7 | 0 | 0 |
| 1 | 0.6 | 1 | 0.8 |
| **164** | **100** | **130** | **100** |

| | | | |
|---|---|---|---|
| 277 | 53.3 | 176 | 60.5 |
| 115 | 22.1 | 72 | 24.7 |
| 22 | 4.2 | 18 | 6.2 |
| 24 | 4.6 | 14 | 4.8 |
| 83 | 16 | 38 | 13.1 |
| 140 | 26.9 | 64 | 22 |
| 21 | 4 | 22 | 7.6 |
| 56 | 10.8 | 40 | 13.7 |
| 27 | 5.2 | 26 | 8.9 |
| 66 | 12.7 | 56 | 19.2 |
| 35 | 6.7 | 23 | 7.9 |
| 12 | 2.3 | 13 | 4.5 |
| 63 | 12.1 | 44 | 15.1 |
| 6 | 1.2 | 12 | 4.1 |
| 5 | 1 | 4 | 1.4 |
| **520** | **100** | **291** | **100** |

| | | | |
|---|---|---|---|
| 22 | 18.8 | 51 | 63 |
| 12 | 10.3 | 19 | 23.5 |
| 7 | 6 | 5 | 6.2 |
| 9 | 7.7 | 3 | 3.7 |
| 11 | 9.4 | 9 | 11.1 |
| 7 | 6 | 10 | 12.3 |
| 38 | 32.5 | 26 | 32.1 |
| **117** | **100** | **81** | **100** |

| | | | |
|---|---|---|---|
| 1,227 | 76.2 | 175 | 41.9 |
| 322 | 20 | 235 | 56.2 |
| 60 | 3.7 | 7 | 1.7 |
| 1 | 0.1 | 1 | 0.2 |
| **1,610** | **100** | **418** | **100** |

| | | | |
|---|---|---|---|
| 506 | 68.2 | 21 | 52.5 |
| 196 | 26.4 | 9 | 22.5 |
| 30 | 4 | 4 | 10 |
| 70 | 9.4 | 9 | 22.5 |

| | | | |
|---|---|---|---|
| 29 | 3.9 | 1 | 2.5 |
| 26 | 3.5 | 1 | 2.5 |
| 36 | 4.9 | 1 | 2.5 |
| 10 | 1.3 | 0 | 0 |
| 4 | 0.5 | 1 | 2.5 |
| 3 | 0.4 | 0 | 0 |
| 6 | 0.8 | 0 | 0 |
| 6 | 0.8 | 2 | 5 |
| 2 | 0.3 | 1 | 2.5 |
| 1 | 0.1 | 0 | 0 |
| 6 | 0.8 | 1 | 2.5 |
| 8 | 1.1 | 0 | 0 |
| 7 | 0.9 | 2 | 5 |
| **742** | **100** | **40** | **100** |
