## Supplemental File 2 for "Effects of a multimedia campaign on HIV self-testing and PrEP outcomes among young people in South Africa: A mixed-methods impact evaluation of ‘MTV Shuga Down South’"

**Individual interviews with young people**

**Time:** 2 hours

**Objective:** To conduct individual interviews with young people who have engaged with MTV Shuga DS2

**Participants:** Young people aged 15-24 living in the Mthatha who have completed the MTV Shuga DS2 questionnaire

**Venue:** A private space – over the phone

**Preparations:** Tablet, smartphone, or laptop to watch scenes, voice recorder, notebook, pencil/pen.

---

**Part 1. How do young people engage with MTV Shuga Down South 2 (DS2)?**

*Explore how MTV Shuga DS2 is experienced by young people (in their own words & experiences)*

- Tell me how you interacted with DS2, through watching the show, listening to the radio, interacting online?
  - How many episodes did you watch or listen to?
- Why did you watch/listen to DS2?
  - What do you like about DS2?
  - How did MTV Shuga make you feel? Did those feelings make you want to engage more or less with the show?
  - What storylines or characters were the most important to you?
  - What kept you engaged in the show?
  - Was there anything that you didn’t like about the show?

- Where did you watch/listen to DS2?
    - When you were watching/ listening, could you describe what was usually going on around you?
    - Did you do any other things while you were watching or listening to DS2. (prompts; doing chores, homework, or talking on the phone)
    - What made it difficult to watch/ listen to the show?
- Did you watch/ listen to DS2 with anyone?
    - What was that experience like to watch it with them?
    - If you watched it on your own, why?
    - Is there anyone with who you wouldn't want to watch/listen to DS2 with? Why?
    - Would you feel comfortable watching/listening to DS2 with a guardian or parent?
- Did you discuss MTV Shuga DS2 with anyone (prompts; friends, family, peers), or did anyone discuss it with you?
    - What did you discuss? Could you give me some examples of scenes or episodes or characters?
    - How did the discussions make you feel?
    - Did you learn or were you influenced by anything in these discussions? *(Probe for examples)*
    - Did people learn from you or did you influence anyone in these discussions? *(Probe for examples)*

- How did you interact with MTV Shuga DS2 besides listening or watching?

  - Did you participate in polls, or post or read conversations online? *(Probe for examples)*
  - What did you like or dislike about interacting online?
- How did discussing with people/interacting with the content online impact your overall experience of Shuga down south 2?
  - Did it change the way you saw characters or their stories? How?
  - Did it make you feel more engaged with the main content? How?
  - Did discussing and engaging with DS2 make you feel supported by the people you discussed with?

***Part 2. If and how DS2 led to change in their lives (based on their memory)***

In this section, we are want to ask you about how Shuga might have changed your behaviour?

***Knowledge / Capability***

- What did you learn about HIV self-screening from the series?
- What did you learn about PrEP (pre-exposure prophylaxis) from the series?

*If they don't remember anything*

- HIVSS: "If you think back to DS2 can you remember the scenes about HIV self-screening? In one of the scenes, Ipeleng and Daniel take a HIV self-test together in her room. In another scene Reggie and Kwanele go to the Taxi Rank and Kwanele privately gets tested.
- PrEP: "If you think back to the show can you remember the scenes about PrEP? One of the scenes Odirile discusses prep with Reggie. In another scene Dineo goes to the clinic and the nurse gives here PrEP."

*If they still don't remember then skip to part 3.*

- Do you trust the information in the show? *(probe why or why not)*

- Do you think the stories about PrEP and HIV self-screening are believable? Would they happen in real life? *(Probe for examples)*
- After watching the show, would you feel confident that you know what HIV self-screening and PrEP are and how they work? *(probe for an explanation)*
    - Did DS2 make you feel more confident about knowing how and where to access PrEP or self-screening? *How did it make you feel more confident? Could you tell me where you could access PrEP?*
    - Did DS2 make you feel more confident in knowing who would benefit from taking PrEP? *(Could you explain that?)*

***Motivation***

Next, we want to ask you about your motivation for accessing HIV self-screening and PrEP

*HIV self-screening.*

If you think back to DS2 can you remember the scenes about HIV self-screening?

- How did the scenes about HIV self-screening make you feel? *(probe for emotions)*
- How are your decisions-and motivations for HIV self-screening different from before you saw the show?
- Did DS2 make you feel more motivated to get tested? Why/why not?
- Have you been tested because of the show?
- Has the show influenced the way you would like to take an HIV test? (i.e. at home, in a clinic, on your own, etc.)

PrEP

Can you remember the scenes about PrEP?

- How did the scenes about PrEP make you feel?

- How are your decisions and motivations for PrEP different from before you saw the show?
- Do you think DS2 makes people more motivated to take or enquire about PrEP? (could you explain?)
- Did DS2 make you think PrEP might be appropriate for you? *If so-* Have you enquired about prep? Why or why not.

**Opportunity**

Next we want to ask you about your opportunity for accessing HIV self-screening and PrEP

- Did DS2 give you an opportunity to discuss (in person or online) things that you wouldn't have talked about before watching the show? Examples?
- Did you have any discussions around HIV self-screening or PrEP?
  - Does DS2 make it easier to talk to your partner about HIV self-screening or PrEP?
  - Does DS2 make it easier to talk to your family about HIV self-screening or PrEP?
- Has DS2 made it easier for you to access PrEP or HIV self-screening? *(Probe beyond motivation and knowledge)*
- Has DS2 made it more difficult for you to access PrEP or HIV self-screening?

**Part 3. Show the DS2 scenes (the edited package):**

**I am now going to send you some clips of the show. You might remember them.**

1. *The scene where Ipeleng & Daniel learn about HIV self-screening, decide to use it, test themselves, and what they do next.*
2. *The scene about Reggie and Kwanele going to get tested in the Taxi Rank.*

3. *The scene about Odirile discussing PrEP with Reggie.*
4. *Dineo's story about PrEP*

- What did you think about these scenes?
- Do you remember seeing these scenes when you watched the show initially?
  - If no, why do you think you don't remember it?
- Did you learn anything new from these scenes?
  - Was the message clear?
- What would you change about these scenes?
  - How could it be better?
- What did you like about these scenes?
- What are your feelings about HIV self-screening and Prep?
- Do these scenes motivate you to use PrEP or Self Screening?

One of the things we want to understand is the elements in the show that make young people want to keep watching the show and what makes them want to tune into the next episode. So, I just want to ask you about certain elements of MTV Shuga, and I want you to tell me how important they were in making you want to engage in the show.

| |
|---|
| The characters |
| The actors |
| The storyline |
| Engagement with MTV<br>(polls, social media) |
| Tailored to SA context |

| |
|---|
| Education value |
| The music |
| Production value |

*End*

- We are now at the end of this interview. Is there anything you want to add before we end?

Thank you so much for answering my question.
