## Supplemental File 3 for "Effects of a multimedia campaign on HIV self-testing and PrEP outcomes among young people in South Africa: A mixed-methods impact evaluation of ‘MTV Shuga Down South’"

**Focus groups with young people**

**Time:** 3 hours (including a 20 min break)

**Objective:** To conduct focus group discussions with young people who have engaged with MTV Shuga DS2

**Participants:** Young people aged 15-25 living in the Mthatha who have complete the MTV Shuga DS2 questionnaire

**Venue:** Over the Zoom

**Preparations:** Voice recorder, mobile phone, Airtime/ Data, Vimeo clips, notebook, ground rules. All participants should have filled out the consent forms before the focus groups.

---

**Part 1. How do young people engage with MTV Shuga Down South 2 (DS2)?**
*Explore how MTV Shuga DS2 is experienced by young people (in their own words & experiences)*

Go around the room and introduce ourselves.

- How many episodes of MTV Shuga did you see and where did you see it?
- What do you like about the series?
- 
- What storylines or characters were the most important to you?
- Was there anything that made it difficult to watch/ listen to the show?
- Did you watch/ listen to DS2 with anyone?
    - Would you feel comfortable watch/listen to DS2 with a guardian or parent?
- Did you discuss DS2 with anyone (i.e. friends, family, peers), or did anyone discuss with you?
    - What did you discuss? Give examples of scenes or episodes, characters.
- Did you participate in polls, or post or read conversations online? Examples.

**Part 2. If and how MTV Shuga DS2 led to change in their lives… (based on their memory)**

**Knowledge / Capability**

- What did you learn about HIV self-screening from the series?

    - Prompt- if they say they didn't learn anything: Do you remember the scenes where they discuss self-screening? Can you describe what happened in those scenes?

- What did you learn about PrEP from the series?
    - Prompt- if they say they didn't learn anything: Do you remember the scenes where they discuss prep? Can you describe what happened in those scenes?

- Do you trust the information in the show?

- Do you think the stories about PrEP and HIV self-screening are believable? Would it happen in real life?  Explain.

**Motivation**

- Did the show change your attitudes of behaviours around HIV self-screening?

- Did the show change your attitudes of behaviours around PrEP different from before you saw the show?

**Part 3. Show the DS2 scenes (the edited package):**

1. *The scene where Ipeleng & Daniel learn about HIVST, decide to use it, test themselves, and what they do next…*
2. *The scene about Reggie and Kwanele going to get tested in the Taxi Rank.*

*Ask all the below questions then send this second*

3. *The scene about Odirile discussing PrEP with Reggie.*
4. *Dineo's story about PrEP*

- What did you think about the scenes?
- Do you remember seeing these scenes when you watched the show initially?
- Did you learn anything new from these scenes?
- What would you change about this scene?
- What are your feelings about HIV self-screening /Prep?

**Part 4**

What did you think about the music in the show?

What did you think about the show being set in a South African context?
