## Supplemental Tables 5-6 for "Effects of a multimedia campaign on HIV self-testing and PrEP outcomes among young people in South Africa: A mixed-methods impact evaluation of ‘MTV Shuga Down South’"

**Table S5**. Mechanisms by which MTV Shuga Down South 2 influenced behaviours through Capability, Motivation and Opportunity to improve HIV self-testing and PrEP outcomes: themes emerging from deductive analysis of qualitative research in Mthatha, Eastern Cape

| Themes *[COM-B* drivers]* | Findings | Sample quotations for HIV Self Screening | Sample quotations for PrEP |
|---|---|---|---|
| Awareness of HIVST & PrEP<br><br>*[Capability]* | • For those who remembered DS2 scenes featuring PrEP and HIV self-testing, awareness and understanding of both increased.<br>• For many, DS2 was the first time they heard about HIV ST and PrEP.<br>• Some participants missed or did not remember relevant scenes or were distracted while viewing and did not gain awareness. | *"Honestly speaking, I didn't know about the self-testing until I watched the show now. I now know that you can test yourself." - Male, 15-19*<br><br>*"Sometimes I'd be on my WhatsApp or cooking. So, during breaks, I'd go to the kitchen and come back." - Female, 15-19* | *"Yoh! I was very shocked that there something called PrEP. Nobody told us about that. I didn't know that you can get pills that can prevent HIV." –Female, 20-24*<br><br>*"I do not know PrEP. I don't remember it from the show." -Female, 15-19* |
| Confidence<br><br>*[Capability & Motivation]* | • Knowledge gained through DS2 about HIV testing and PrEP, and storylines about people living and thriving with HIV, reduced the fear of knowing their HIV status.<br>• Some participants gained confidence from DS2 to enquire and learn more about HIV ST and PrEP. Some were more confident about enquiring about HIV ST than PrEP as they were already familiar with screening and testing for HIV, whereas PrEP was an innovation that they had never heard of before. | *"I saw how Bongi [a DS2 character who learns she is HIV positive] accepted herself. So, knowing [my status] won't mean the end of the world, I'll just know." - Female, 15-19*<br><br>*"I know what to say when I go get it [HIV self-screening and PrEP], and I know what will be given to me. So, it [DS2] made it easier, because I actually understand what I will be experiencing and the whole situation." – Male, 20-24*<br><br>*"They say HIV is something that is tested through blood so, I kind of don't understand like what they are doing there [with HIV self-* | *"It [DS2] made me want to enquire. I also did a google search on PrEP. I didn't go, you know, like to the health institution to find out, but google-searched it just to know how safe it is and side effects, stuff like that. Just general information about Prep and how to use it." -Male, 20-24*<br><br>*"I would not really be confident talking to people because I don't have a personal experience with it [PrEP], unlike HIV testing. It would be a lot easier to talk about HIV self-testing than PrEP." – Female, 15-19* |

| | | | |
|---|---|---|---|
| | • A few participants said they lacked confidence in HIV ST and PrEP because they were confused by scenes in *MTV Shuga*. | *screening]. Cause there is only saliva inside the mouth, so I don't understand."- Male 15-19* | |
| Reflection<br><br>*[Motivation]* | • Watching different scenarios with MTV Shuga characters using PrEP and HIV ST helped participants reflect on their preferences for using these tools at different times of their life.<br>• For some, the show generated interest using HIV ST because ST is convenient, private, and uses saliva only.<br>• Though some participates were worried about the side effects of PrEP, most were excited to learn about a new way to protect themselves against HIV.<br>• A few participants decided after watching the show that HIV ST / PrEP was not appropriate for them and they would prefer to use other HIV testing and prevention options. | *"I didn't know it [HIV self-testing] existed before I watched the show. I also learnt that it is easy. Listen, some people are scared of needles, and it becomes their reason for not wanting to test. They will be like 'no I'm afraid of needles' so now it's very easy, you just swab in your mouth, then it's done."- Female, 20-24*<br><br>*"After watching the show, I prefer [testing] at home, because if you can't cope, you can even have a friend there. Testing at a clinic would make me feel anxious." – Female, 15-19*<br><br>*"This thing of testing at home really scared me a lot. I prefer to go to the clinic, because from the clinic, if you find yourself positive, you will get counselling at the same time." – Male, 20-24* | *"The show teaches people about PrEP. It's appropriate for me too, because I will be faced with things in my life, so knowing that there is PrEP, at least I'll be safe." – Female, 15-19*<br><br>*"I think it [DS2] shows that when you're with someone, you shouldn't trust that person so much with your life. It's like with Dineo; she tells her partner how important it is to take PrEP and to use a condom regardless of whether you are in a committed relationship. You shouldn't trust your partner so much so that you don't use protection." -Female, 20-24*<br><br>*"I do prefer to use PrEP, but I do not like the side effects of it. But I also like the fact that it protects you from getting HIV when your partner has tested positive." – Female, 15-19* |
| Preparation<br><br>*[Motivation]* | • The series helped participants, some of whom were not currently sexually active, to plan for scenarios and relationships where there they might use PrEP and HIV ST to protect themselves and their partners against HIV in the future. | *"I was not actually active before I watched it, so after I watched it, then I became sexually active, then that's when I felt like going for HIV screening, without having to go like, [to the] doctors. You know? Like, I realised that I actually should know my status."– Male, 15-19* | *"Life is life. Things do happen, so now that I have information about PrEP, it will help me when I meet someone. I will know what I need to do and know how PrEP will help me." -Male, 20-24* |

| | | | |
|---|---|---|---|
| | | *"It [the show] made me feel more motivated to get tested because I do have a partner. I'm just motivated for us to test together. It made me feel confident to be open about my status."*<br>*– Female, 20-24* | *"I now know that if I slept with someone who has HIV, it wouldn't be the end of the world; I could use PrEP to prevent me from getting the virus. Before seeing the show, PrEP was just something I saw in books. I didn't know that it is actually out there. But after seeing the show, I see that it is something that actually helps a lot of people." Female, 20-24* |
| Access to services<br><br>*[Capability & Opportunity]* | • The series made some participants more aware of where they might access HIV ST and PrEP.<br>• However, some doubted that they could access these services, or information about PrEP in Mthatha (where participants were based), which they considered to have different resources than Johannesburg (*slang: Joburg*) the setting of DS2. | *"I watched the show and saw how to get it [a self-test kit], but I wouldn't be confident that if I go to the hospital, I'd get them. The hospitals here are not the same as those in Joburg."*<br>*- Female, 15-19*<br><br>*"I think most clinics are closed right now due to this COVID-19 thing" – Female, 15-19* | *"I will go to the clinic. It's where most people go to in the show Shuga. So, I'll go to a local clinic and consult with a nurse or whichever doctor there is and ask them how can I get PrEP. I believe they will explain what the side effects are." -Male, 20-24*<br><br>*"The nurses at our school don't even know about it [PrEP]. Even when you getting HIV test, and ask about this PrEP, they don't know about it." - Male, 20-24* |

*COM-B: Capability, Opportunity, Motivation [reference]

**Table S6**. Evidence of broader social mechanisms by which MTV Shuga DS2 influenced HIV self-testing and PrEP outcomes: themes emerging from open code analysis of qualitative research in Mthatha, Eastern Cape

| Themes | Findings | Sample quotations |
|---|---|---|
| DS2 inspires and facilitates supportive conversations about sex and HIV | • Participants shared that young people (their peer group) often avoid or find it difficult to talk about sexual health, especially with partners and parents.<br>• Watching the show with friends, partners and parents made it easier to initiate discussions around sexual health, including PrEP and ST.<br>• Discussions about the series triggered open dialogue about young people's sexual relationships, their individual needs and preferences, moral beliefs and decision-making.<br>• Participants felt supported after engaging in conversations about DS2 with others. Some participants said if they need support and advice in the future, they could seek it from the person[s] with whom they watched DS2.<br>• However, some participants did not want to watch DS2 with others, especially parents, because they felt uncomfortable, embarrassed or worried about how they might react to the sexual content. | *"It did [make me want to discuss HIV and PrEP with my partner]. Us, as youth, we tend to duck conversations especially important conversations. So, if it's playing on TV, we would actually want to discuss it." -Female, 20-24*<br><br>*"Our parents do not teach us about HIV, pregnancy, when we start engaging with sexual intercourse. So, I learnt a lot about those things from MTV Shuga, because our parents run away from those things. I would like to watch the show with my mother so that she can learn that being honest and talking about these things will make us more aware of what is going on." - Female, 20-24*<br><br>*"I personally, generally observed a paradigm shift from speaking about alcohol, girls, sex, and hangovers to speaking about things like contraceptive methods, how we can organise PrEP, and how we can expand the knowledge we got from the show. So you assisted us to tap into a different perspective of things we would not have spoken about had we not watched the show." - Male, 20-24*<br><br>*"After watching the show, my partner and I discussed the importance of knowing about each other's statuses. Also, that if he is not being honest and has other girlfriends, he needs to take PrEP so that we are protected." -Female, 20-24*<br><br>*"I saw how my friends were [when watching the show] and that God forbid I would find myself in such a situation where I am HIV positive, I know that I can tell my friends, and they will be very supportive and will make jokes about it and they'll always be there." - Female, 15-19*<br><br>*"My friend and I would discuss specific scenes […] and the characters reactions during the show. It made me feel like I can ask advice from her and also that we can help each other when in certain situations." – Female, 20-24* |

| | | |
|---|---|---|
| | | *"I can't be comfortable [watching the show with my parents] because I get shy with some of the things that are happening there, especially if there are people around me. I am a shy person. Not because… Like, it's alright that they watch, but when they watch, they must alone, not with me, they must do it by themselves, and I will watch it alone..." – Male, 20-24* |
| DS2 viewers wanted DS2 and its content on HIVST and PrEP to be more widespread | • Because many participants learned about PrEP and HIV ST for the first time from DS2, they felt that more people in their community should know about these resources (PrEP in particular). | *"Yes, Dineo going to the clinic to talk about PrEP with the nurse. That is when I got to realise that there was something called PrEP because I didn't know about it. […] I feel like [The Department of Health] should be motivating the youth to take part in it. Like I said, I've never heard about it before." - Female, 15-19*<br><br>*"I feel like people are not really informed about it [PrEP] and that they need to know about it in order to be able to use it." - Female, 20-24*<br><br>*"I just wish that lot of young people could watch the show. I just wish a lot of people will know about the PrEP, know about the HIV self-screening." -Female, 15-19*<br><br>*"They [HIV SS and PrEP]are both important, but in terms of PrEP, there should be more awareness campaigns, especially in the rural areas. – Female, 20-24* |
| DS2 viewers become advocates for HIV testing & prevention | • Knowledge and confidence gained from MTV Shuga about HIV prevention and testing emboldened some young people to share and educate friends, family and partners about HIV ST and PrEP and engaging in safe sex. | *"It [the show] made me aware and able to talk about HIV self-screening. Yes, it's something that I can confidently talk about only because of the show now. Because people are usually afraid of the pain and pricking when it comes to taking an HIV test. So, I'll definitely be an advocate for the self-screening HIV test now because of the show."- Male, 20-24*<br><br>*"The show made me feel like I could share information with others, especially on preventing risky issues from not happening. I could give knowledge with regards to that." - Female, 15-19*<br><br>*"Now that I know more about PrEP from the show, I would suggest it to my girl or my partner, whoever I am having, an encounter with." – Male, 15-19* |

| | | |
|---|---|---|
| | | *"I can be able to give my little brother advice on when he wants to date. So, now I can [after watching the show] explain to him how to be safe when he's having sex, testing, everything." – Male, 20-24* |
