## Supplementary figures and images for "Effects of a multimedia campaign on HIV self-testing and PrEP outcomes among young people in South Africa: A mixed-methods impact evaluation of ‘MTV Shuga Down South’"

### Supplemental Figure 1

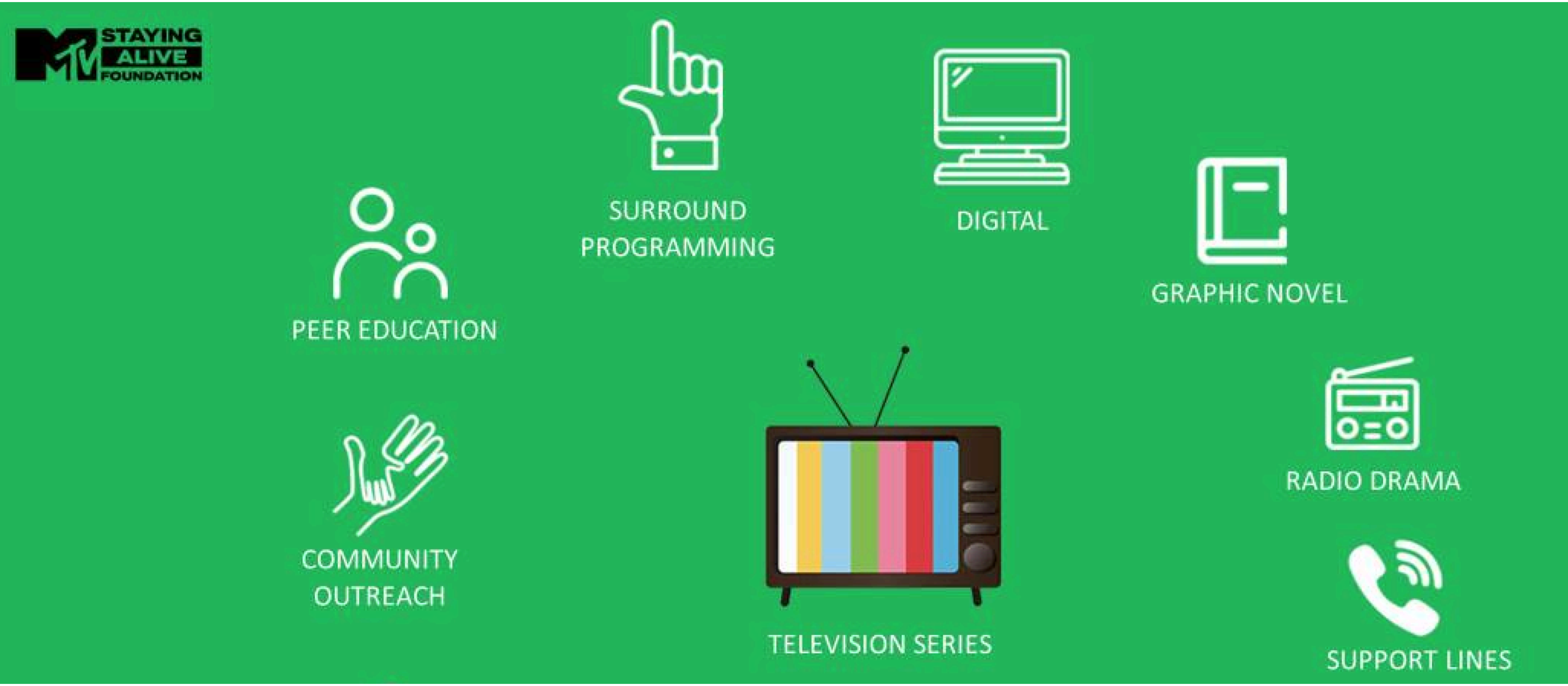

### Supplemental Figure 2

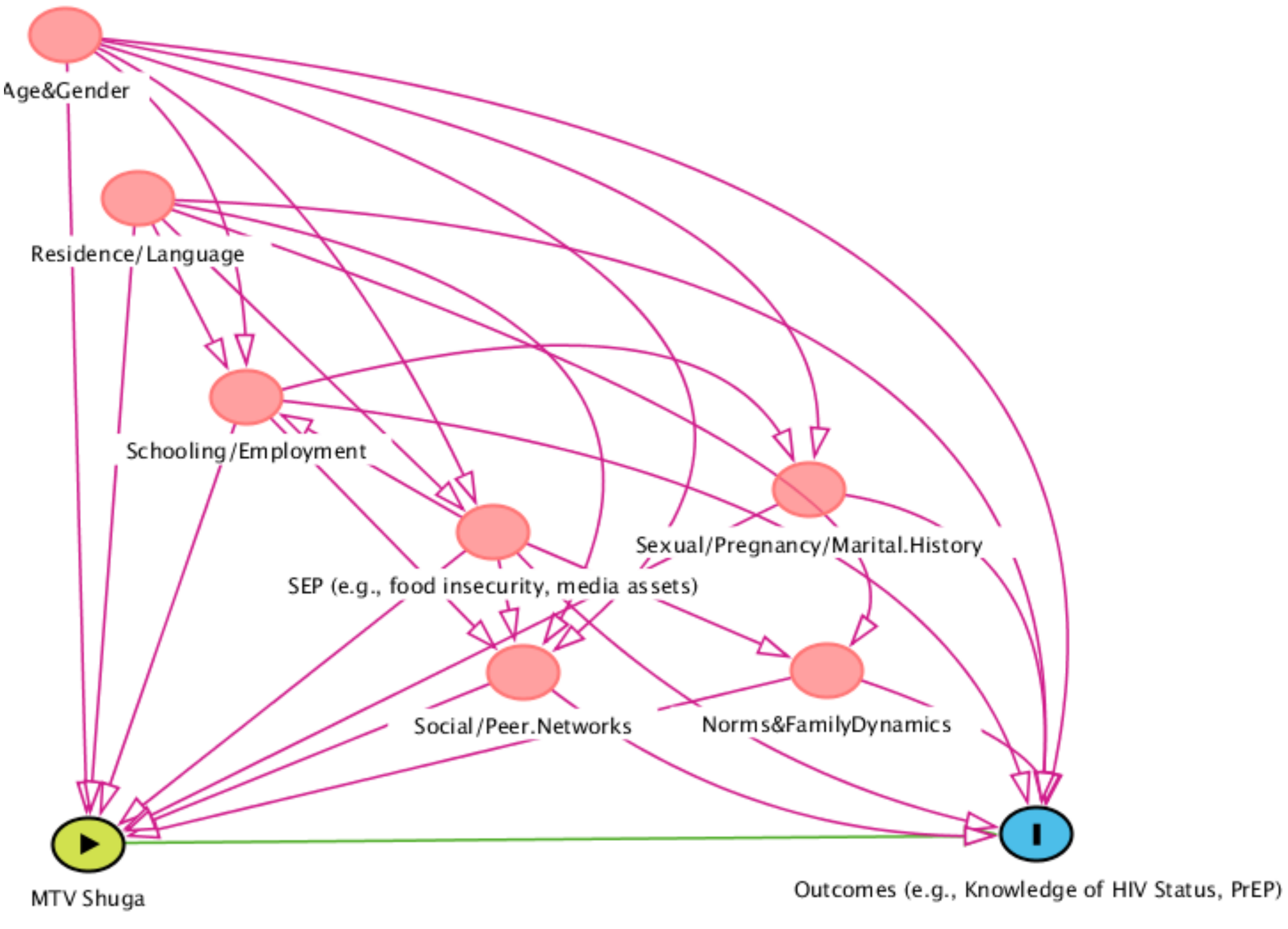
